# A rare variant in D-amino acid oxidase implicates NMDA receptor signaling and cerebellar gene networks in risk for bipolar disorder

**DOI:** 10.1101/2021.06.02.21258261

**Authors:** Naushaba Hasin, Lace M. Riggs, Tatyana Shekhtman, Justin Ashworth, Robert Lease, Rediet T. Oshone, Elizabeth M. Humphries, Judith A. Badner, Pippa A. Thompson, David C. Glahn, David W. Craig, Howard J. Edenberg, Elliot S. Gershon, Francis J. McMahon, John I. Nurnberger, Peter P. Zandi, John R. Kelsoe, Jared C. Roach, Todd D. Gould, Seth A. Ament

## Abstract

Bipolar disorder is an often-severe mental health disorder characterized by alternation between extreme mood states of mania and depression. Despite strong heritability and the recent identification of 64 loci of small effect, pathophysiological mechanisms and much of the genetic risk remain unknown. Here, through genome sequencing and linkage and association analyses, we found that rare variants co-segregating with bipolar disorder in large multiply affected families cluster within gene networks enriched for synaptic and nuclear functions. The top variant in this analysis prioritized by statistical association, predicted deleteriousness, and network centrality was a missense variant in the gene encoding *D*-amino acid oxidase (*DAO*^*G131V*^). Heterologous expression of DAO^G131V^ in human cells resulted in decreased DAO protein abundance and enzymatic activity. In a knock-in mouse harboring this human *DAO*^*G131V*^ variant, *Dao*^*G130V/+*^, we similarly found decreased DAO protein abundance, as well as enhanced stress susceptibility and blunted behavioral responses to pharmacological inhibition of *N*-methyl-D-aspartate receptors (NMDARs). RNA sequencing of cerebellar tissue revealed that *Dao*^*G130V*^ resulted in decreased expression of two gene networks that are enriched for synaptic functions and for genes expressed specifically in Purkinje cells and granule neurons. Similar expression changes in both of these gene networks were also identified in the cerebellum of bipolar disorder cases vs. controls. These findings implicate dysregulation of NMDAR signaling and of gene expression in cerebellar neurons in bipolar disorder pathophysiology and provide insight into its genetic architecture.

**One Sentence Summary:** Functional studies of a rare hypofunctional variant in the D-amino acid oxidase gene implicate stress susceptibility, NMDA receptor signaling, and cerebellar circuits in risk for bipolar disorder.

## INTRODUCTION

Bipolar disorder is a severe mental health disorder affecting 1-2% of the population and characterized by alternation between extreme mood states of mania and depression. Bipolar disorder is strongly familial, with 8 to 10-fold relative risk in the first-degree relatives of probands, ∼70% concordance among monozygotic twins, and broad-sense heritability in the range of 60-80% *(1)*. While genome-wide association studies (GWAS) of common variants have identified 64 risk loci for bipolar disorder, causal mechanisms remain elusive due to the small effect sizes of common risk variants and their predominant localization to non-coding regions of the genome *(2, 3)*.

Identifying rare, larger-effect risk variants would aid in the discovery of disease mechanisms, development of more precise disease models, and advancement of novel therapeutic targets. Several studies have reported rare variants that co-segregate with bipolar disorder in large, multiply affected pedigrees. These variants were enriched in a variety of functional categories related to neuronal functions and suggested overlap with risk genes for related psychiatric disorders such as schizophrenia and autism spectrum disorder *(4–8)*. Enrichments in similar functional categories have been detected by exome sequencing of larger case-control cohorts *(9, 10)*. However, both family studies and case-control cohorts indicate extensive heterogeneity and polygenicity. Underlying mechanisms remain unclear since very few of these variants have been functionally characterized *(7, 11)*.

Here, we applied network analysis and multi-parameter optimization to prioritize specific variants in pedigrees multiply affected with bipolar disorder. The top-scoring variant was a missense variant in the *DAO* gene. *DAO* encodes the *D*-amino acid oxidase (DAO) enzyme, which catalyzes the oxidative deamination of *D*-amino acids, including the requisite *N*-methyl-D-aspartic acid receptor (NMDAR) co-agonist *D-*serine. We functionally characterized this variant in human cells and in a genetically precise knock-in mouse to gain insight into its molecular, cellular, and behavioral consequences.

## RESULTS

### A rare, nonsynonymous variant in the *DAO* gene associated with risk for mood disorders

We studied whole-genome sequences (WGS) of 200 individuals in 41 multiply affected pedigrees to identify rare variants that co-segregate with bipolar disorder. Previously, we reported a candidate gene-based analysis of these sequences *(5)*. Here, we extended our analysis to all protein-coding genes. We annotated rare and uncommon single-nucleotide variants (MAF < 5% in the 1000 Genomes Phase 3 reference panel) with a predicted functional effect on a protein-coding gene, including protein-truncating variants, missense variants, splice site variants, and non-coding variants in a gene’s 5’ or 3’ untranslated regions. We selected variants with evidence of genetic linkage or association to bipolar disorder by two methods. First, we tested for genetic associations using linear mixed models. Although this analysis is underpowered to detect associations at a conventional threshold for genome-wide significance, we identified 684 functional variants nominally associated with an increased risk for bipolar disorder (*P* < 0.05). In addition, we used our WGS to fine-map previously determined regions of suggestive linkage (per-pedigree LOD scores > 0.6, *P* < 0.05), based on genotyping of 4,500 genome-wide single nucleotide polymorphisms (SNPs) in a larger number of individuals in the same 41 pedigrees *(12)*. This strategy identified 168 additional variants. In total, we identified 852 variants in 741 genes (Fig. 1A; Table S1). Of these, 12 were annotated as protein-truncating variants, seven as splice site variants, 348 as missense variants, and 527 as non-coding.

**Figure 1.**
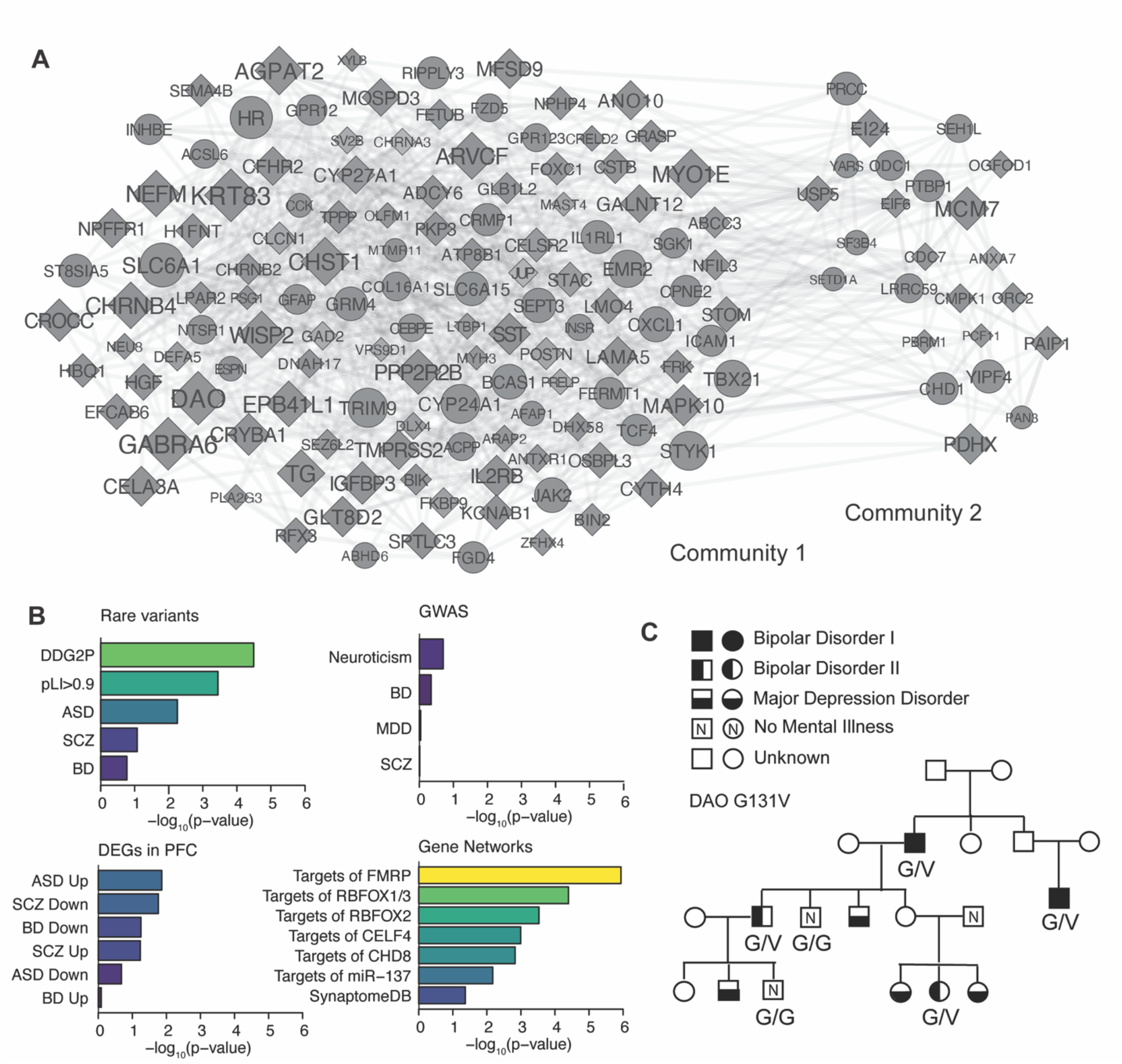
Identification of functional variants in multiply affected pedigrees with bipolar disorder. A. Contextualization of the top 150 genes identified in bipolar disorder pedigrees in a protein interaction network. Node shape: diamond = coding; circle = non-coding. Node color: p-value for linkage or association (darkest red corresponds to *P* = 1.0e-5). Node size: ranking on combined evidence from statistical association, deleteriousness, and network centrality. B. Enrichments for neuropsychiatric risk genes and related gene sets: rare variants = risk genes from exome and genome sequencing studies; GWAS = genes at GWAS risk loci; DEGs in PFC = Differentially expressed genes in prefrontal cortex; gene networks. C. *DAO*^*G131V*^ co-segregated with bipolar disorder in a four-generation pedigree.

Exome and genome sequencing studies of adult-onset psychiatric disorders have identified only a handful of high-confidence risk genes. We tested for overlap of the 741 genes identified in our bipolar disorder pedigrees with known risk genes for psychiatric disorders and related traits (Fig. 1B; Table S2; Supplementary Materials and Methods). Consistent with findings from other neuropsychiatric disorders *(13, 14)*, we found that 138 of the 741 genes are loss-of-function intolerant, suggesting that their complete loss-of-function reduces fitness (over-representation of genes with gnomAD pLI > 0.9: odds ratio = 1.41, *P* = 3.6e-4). Mutations in 108 of the 741 genes are associated with severe developmental disorders in the Developmental Disorder Genotype-Phenotype Database *(15)* (odds ratio = 1.6, *P* = 3.2e-5), including 10 of 102 high-confidence risk genes for autism spectrum disorder from large-scale exome sequencing by the Autism Sequencing Consortium *(16)*: *RFX3, DYRK1A, SETD5, SLC6A1, DIP2A, TCF4, GFAP, PRR12, PPP1R9B*, and *NSD1* (odds ratio = 2.8, *P* = 5.5e-3). Intersecting our gene list with results from the largest available exome sequencing datasets of psychiatric disorders revealed that 49 of our 741 genes were nominally associated with risk for schizophrenia in the SCHEMA dataset *(17)* (odds ratio = 1.3, *P* = 0.08), including the top schizophrenia risk gene, *SETD1A*. Seven of our 741 genes were nominally associated with risk for bipolar disorder in the BipEx dataset *(10)*, which does not include individuals from these 41 pedigrees: *PPIL2, TMEM156, SBSPON, SEMA4B, CHD1L, CHRNB2*, and *SCLY* (odds ratio = 1.6, *P* = 0.17).

We also tested for overlap with genes and gene networks that have been implicated in neuropsychiatric disorders by other approaches. Perhaps surprisingly, we found little overlap of our 741 genes with genes proximal to risk loci from genome-wide association studies (GWAS) of bipolar disorder *(2)*, major depression *(18)*, schizophrenia *(19)*, or neuroticism *(20)*. We found modest overlap with genes differentially expressed in the prefrontal cortex of individuals with schizophrenia, autism, and bipolar disorder *(21)*. We found strong overlap with several gene networks implicated in risk for multiple neuropsychiatric disorders *(22)*, including targets of the neuronal RNA binding proteins FMRP, RBFOX2, RBFOX1/3, and CELF4, targets of microRNA-137, and targets of the chromatin-remodeling protein CHD8.

We contextualized the 741 genes in a protein-protein interaction (PPI) network to gain insight into their functional relationships. We found a significantly higher rate of PPI among our 741 genes than expected, based on interactions from the GeneMania Database *(23)* (*P* < 0.001 based on 1,000 permutations of network edges). Leiden clustering *(24)* revealed that the 741 genes cluster into two major communities (Fig. 1A; Table S3). Community 1 contained 382 genes and was enriched for the Gene Ontology term “synapse” (48 genes, odds ratio = 2.0, *P* = 2.2e-5; Table S4). Community 2 contained 163 genes and was enriched for genes localized to the “nucleus” (163 genes, odds ratio = 2.2, *P* = 1.5e-11; Table S4), including genes involved in “DNA repair” (25 genes, odds ratio = 3.4, *P* = 5.2e-7), “mitotic cell cycle” (36 genes, odds ratio = 2.7, *P* = 9.4e-7), and “chromatin remodeling” (11 genes, odds ratio = 3.9, *P* = 2.2e-4). All of these categories are also enriched among risk genes for related psychiatric disorders, including autism and schizophrenia *(17, 25, 26)*.

We prioritized specific variants based on a combination of network centrality (eigencentrality within each community), the strength of statistical association to bipolar disorder from our linkage and association analyses, and predicted deleteriousness (CADD score *(27)*; Table S3). The top-scoring variant overall was rs768676371, a missense variant in *DAO*, encoding *D*-amino acid oxidase (Fig. 1C). Other top-scoring variants in Community 1 included rs3811993, a missense variant in *GABRA6*, encoding the GABA_A_ receptor α-6 subunit; rs12914008, a missense variant in *CHRNB4*, encoding the cholinergic receptor nicotinic β-4 subunit; and rs41276505, located in the 3’ UTR of *SLC6A1*, encoding GABA Transporter 1. Top-scoring variants in Community 2 included rs1263034966, a missense variant in *MCM7*, encoding Minichromosome Maintenance Complex Component 7; chr11:125452286 C/T, a novel missense variant in *EI24*, encoding Etoposide Induced 2.4, an autophagy-associated transmembrane protein; and rs2455425, located in the 3’ UTR of *CHD1*, encoding Chromodomain-Helicase DNA-binding 1. These genes have well-established functions at synapses and in DNA repair and chromatin remodeling, consistent with the broader enrichments of each community.

DAO catalyzes the degradation of *D*-amino acids including *D*-serine, a well-characterized co-agonist of NMDARs *(28–30)*. In mice, DAO loss-of-function caused elevated *D*-serine levels, increased NMDAR activity, and altered cognition and behavior *(31–34)*. Several previous reports described linkage of the *DAO* region to bipolar disorder and schizophrenia *(35– 40)*. However, common variants in this region have not been shown to be associated with neuropsychiatric disorders. The rs768676371 variant results in a glycine-to-valine substitution at amino acid 131 (henceforth, *DAO*^*G131V*^). This variant was initially discovered under the top previously determined linkage peak *(12)* in the proband of a four-generation pedigree. Using TaqMan genotyping, we confirmed the *DAO*^*G131V*^ genotype in the proband, as well as in three additional members of the family, all of whom had bipolar disorder type-1 or type-2. We also confirmed that the variant was absent in two unaffected family members. Non-parametric linkage analysis of these confirmed genotypes suggested linkage to bipolar disorder within this family (*P* = 7e-4). Replication analysis using exome sequences from the SCHEMA and BipEx consortia *(10, 17)* (*n* = 24,248 schizophrenia cases, 13,933 bipolar disorder cases, and 97,322 non-diseases controls) revealed one additional copy of *DAO*^*G131V*^ in a schizophrenia case and none in controls. In addition, the gnomAD v2.1.1 dataset contains one copy of the allele in an individual from the “non-neuro” cohort (*n* = 104,064) whose mental health status is unknown. Thus, *DAO*^*G131V*^ is a rare variant that we have identified exclusively or near-exclusively in individuals with bipolar disorder or schizophrenia.

### Effects of *DAO*^*G131V*^ on DAO expression and activity

We set out to test the functional consequences of the DAO^G131V^ missense variant. Examination of DAO crystal structures from the Protein Data Bank indicated that G131 is located in a loop domain (Fig. 2A). Protein folding simulations with Rosetta *(41)* substituting glycine with the bulkier valine residue resulted in a predicted increase in the free energy of the folded protein (i.e., destabilization, *P* = 0.004; Fig. 2B). Thus, the model predicts that DAO^G131V^ may cause hypofunction by destabilizing the DAO protein. We tested this prediction via heterologous expression of DAO^G131V^ or non-mutant DAO protein in the HEK293T cell line, which does not natively express *DAO*. Quantitative PCR (qPCR) demonstrated similar levels of *DAO*^*G131V*^ vs. non-mutant *DAO* transcripts, suggesting that *DAO*^*G131V*^ does not influence transcription or mRNA stability (Fig. S1A). By contrast, quantification of DAO protein in transfected HEK293T cells by Western blot revealed ∼30% lower levels of DAO^G131V^ vs. non-mutant DAO protein (*F* = 5.7, *P* = 0.017, Fig. 2C,D; Fig. S1B-D). DAO^G131V^ did not alter the subcellular localization of the DAO protein in transiently transfected HEK293T cells (Fig. S1E,F). These results suggest that DAO^G131V^ leads to reduced abundance of the DAO protein, possibly via decreased protein stability. DAO^G131V^ may also inhibit formation of the active DAO homodimer, as the variant is located at the dimerization interface *(42)*.

**Figure 2.**
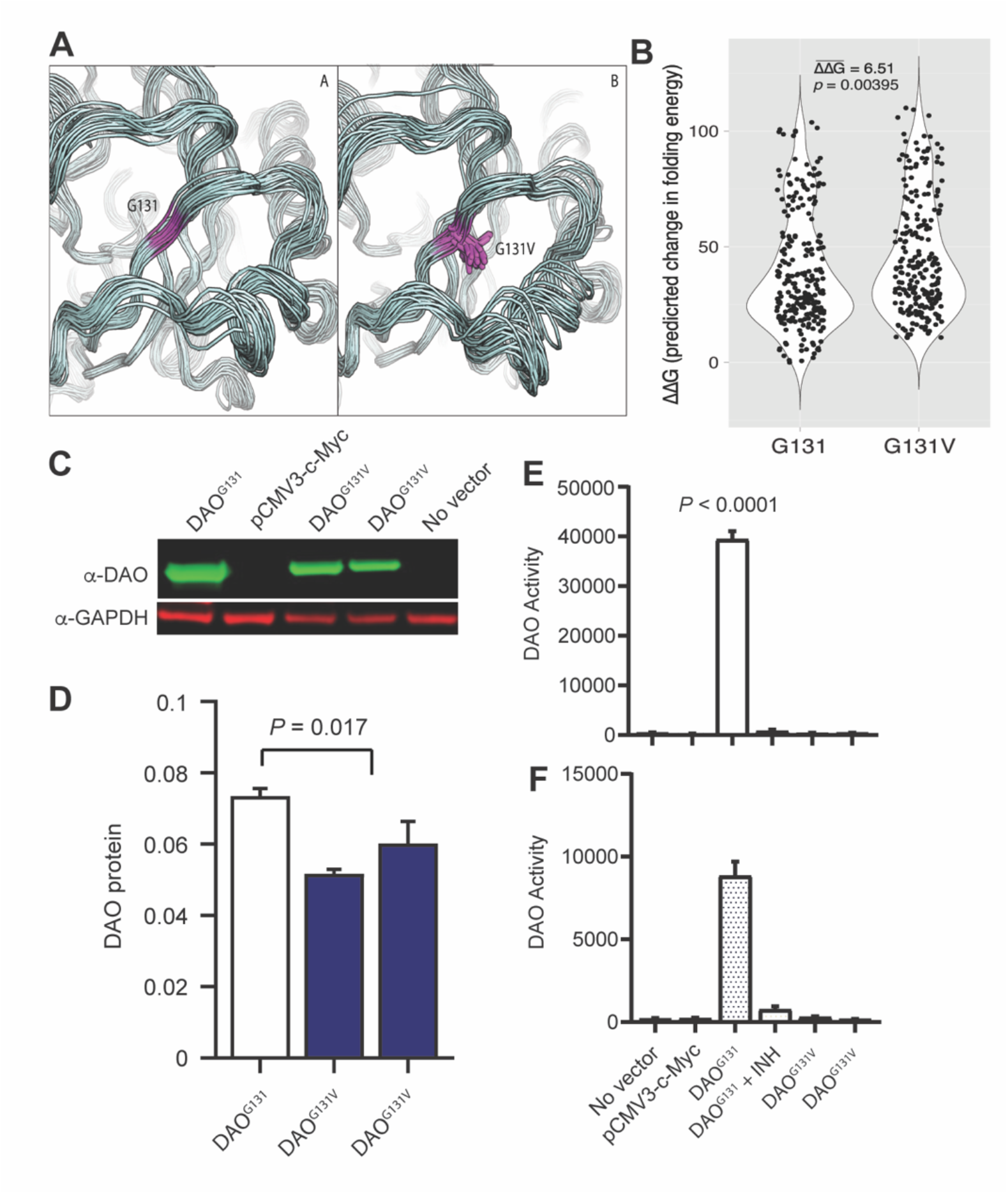
*DAO*^*G131V*^ results in hypofunction of the DAO protein in human cells. A. Structural modeling of the loop region of the DAO protein in the presence of the reference G131 allele (left) and with G131V (right). B. Free energy predictions from protein folding simulations of the DAO protein with G131 vs. G131V. C. Western blot showing DAO protein abundance in HEK293T cells following transfection with DAO^G131^ or with DAO^G131V^. D. Quantitation of DAO protein with G131 vs. G131V. E,F. Effects of DAO^G131V^ on DAO enzymatic activity in HEK293T cells in the presence (E) or absence (F) of the co-enzyme FAD.

Next, we investigated the effect of DAO^G131V^ on DAO enzymatic activity. The *D*-amino acid oxidase reaction produces hydrogen peroxide (H_2_O_2_) as a biproduct. Therefore, DAO activity is commonly quantified using Amplex red assays, which directly measure H_2_O_2_ production *(43)*. We detected robust H_2_O_2_ production in cell lysates from HEK293T cells expressing non-mutant DAO in the presence of *D*-serine substrate (Fig. 2E,F). H_2_O_2_ production was blocked in the presence of the DAO inhibitor 6-methyl-benzo[d]isoxazole-3-ol, confirming that this signal corresponds to DAO activity. By contrast, DAO activity was essentially undetectable in cells expressing DAO^G131V^. Similar results were obtained with (*P* < 1.0e-4, Fig. 2E) or without (*P* < 1.0e-4, Fig. 2F) addition of 10 μM FAD co-enzyme. Therefore, DAO^G131V^ results in almost complete loss of DAO activity in HEK293T cells.

The human DAO protein shares 80-85% homology across higher vertebrates and is present in most eukaryote and prokaryote genomes. Multiple sequence alignment of the DAO protein revealed that the G131 residue is conserved across all examined vertebrate and invertebrate species (Fig. S2A). Thus, DAO G131 is a deeply conserved residue in the DAO protein. To investigate the effects of DAO^G131V^ *in vivo*, we generated a genetically precise knock-in mouse model. CRISPR/Cas9 genome editing was used to introduce a G-to-V point mutation into the germline of C57BL/6N mice at the homologous position in the *Dao* gene (Fig. S2B-E). This position in offset by one amino acid in the human vs. mouse protein. Therefore, we refer to the resulting mouse allele as *Dao*^*G130V*^. Successful editing of four heterozygous *Dao*^*G130V/+*^ founders was confirmed by sequencing, as well as by a restriction fragment length polymorphism assay.

We assessed the effects of *Dao*^*G130V*^ on RNA expression and protein abundance in several mouse tissues by qPCR and Western blots. Previous studies have demonstrated high levels of *DAO* mRNA and protein primarily in astrocytes of the mouse hindbrain *(44)*, though the activity of DAO in the human brain may be broader *(45)*. High expression is also observed in the kidney. As expected, *Dao* mRNA (Fig. 3A) and DAO protein (Fig. 3B,C) were most abundant in kidney, followed by two hindbrain regions, cerebellum and medulla. Expression of *Dao* mRNA and DAO protein were very low in both forebrain regions tested, hippocampus and cortex. *Dao* mRNA expression in each tissue was similar in *Dao*^*G130V/+*^ heterozygotes and *Dao*^*G130V/G130V*^ homozygotes vs. wildtype *Dao*^*+/+*^ mice (Fig. 3A), suggesting that the variant has little effect on transcription or mRNA stability. By contrast, *Dao*^*G130V*^ caused a substantial dose-dependent reduction in the DAO protein level, which was most pronounced in the cerebellum (*F* = 21.8, *P* = 3.0e-6), medulla (*F* = 22.8, *P* = 1.8e-6), and kidney (*F* = 14.6, *P* = 1.4e-4, Fig. 3B,C). Therefore, *in vivo Dao*^*G130V*^ results in reduced levels of DAO protein, consistent with our results in human cells.

**Figure 3.**
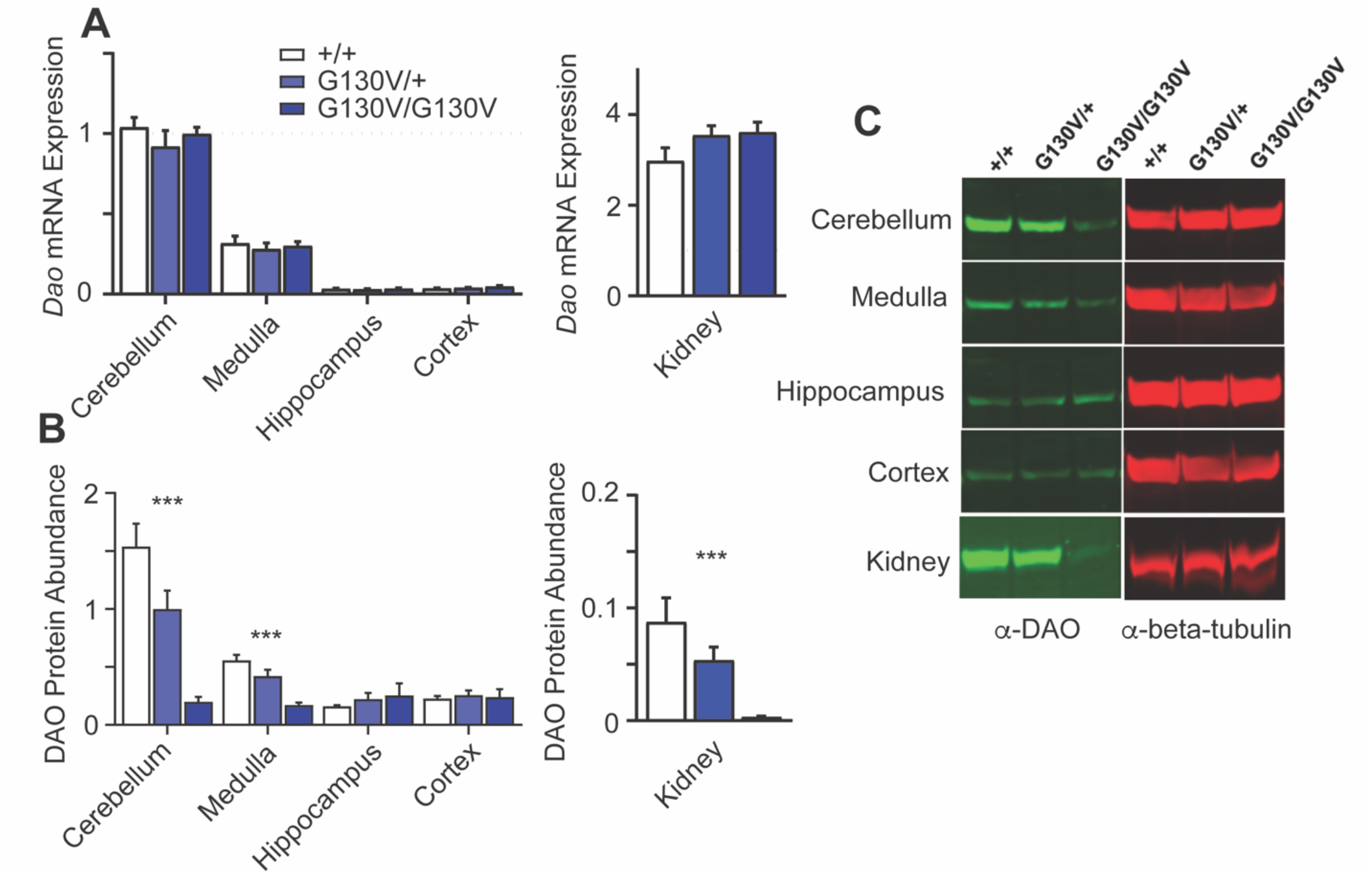
*Dao*^*G130V*^ results in reduced DAO protein abundance in mouse tissues. A. Expression of *Dao* mRNA in brain regions (left) and kidney (right) from Dao^+/+^, Dao^G130V/+^, and Dao^G130V/G130V^ mice (n=5 / genotype). B,C. Abundance of DAO protein in brain regions (left) and kidney (right) from Dao^+/+^, Dao^G130V/+^, and Dao^G130V/G130V^ mice (n=5 / genotype). *** *P* < 0.001

### In vivo behavioral characterization of the *Dao*^*G130V*^ variant in knock-in mice

We performed a sequence of experiments to assess the impact of the *Dao*^*G130V*^ variant on behavioral responses *in vivo*. Previous studies have demonstrated that mice homozygous for DAO loss-of-function mutations manifest behavioral changes relevant to mood and psychotic disorders, including increased anxiety-like behaviors *(46, 47)*. However, behavioral phenotypes in heterozygous mice with DAO mutations have not been reported. Given that we observed only heterozygous *Dao*^*G131V*^ genotypes in humans, we therefore characterized the behavioral phenotypes of heterozygous *Dao*^*G130V/+*^ mice vs. wildtype *Dao*^*+/+*^ controls.

First, we assessed general locomotor activity and basal anxiety-like responses in the open field test and elevated plus maze. Both *Dao*^*G130V/+*^ and wildtype mice displayed normal habituation to the novelty of the open field environment, with no effect of genotype on distance traveled either in males or females (males: F_(1, 21)_ = 0.99, *P* > 0.1; females: F_(1, 22)_ = 0.01, *P* > 0.1; Fig. 4A). Also, *Dao*^*G130V/+*^ and *Dao*^*+/+*^ mice responded similarly to the anxiogenic properties of the open field, as all groups spent a similar amount of time in the center zone of the arena (*P* > 0.1, Fig. S3A). Similarly, neither sex (*F*_(1, 43)_ = 2.5, *P* > 0.1) nor genotype (*F*_(1, 43)_ = 0.0, *P* > 0.1) influenced the amount of time spent in the open arms of the elevated plus maze (Fig. S3B). These results suggest that in heterozygous mice *Dao*^*G130V*^ does not alter general locomotor activity and does not promote an anxiety-like phenotype in these assays.

**Figure 4.**
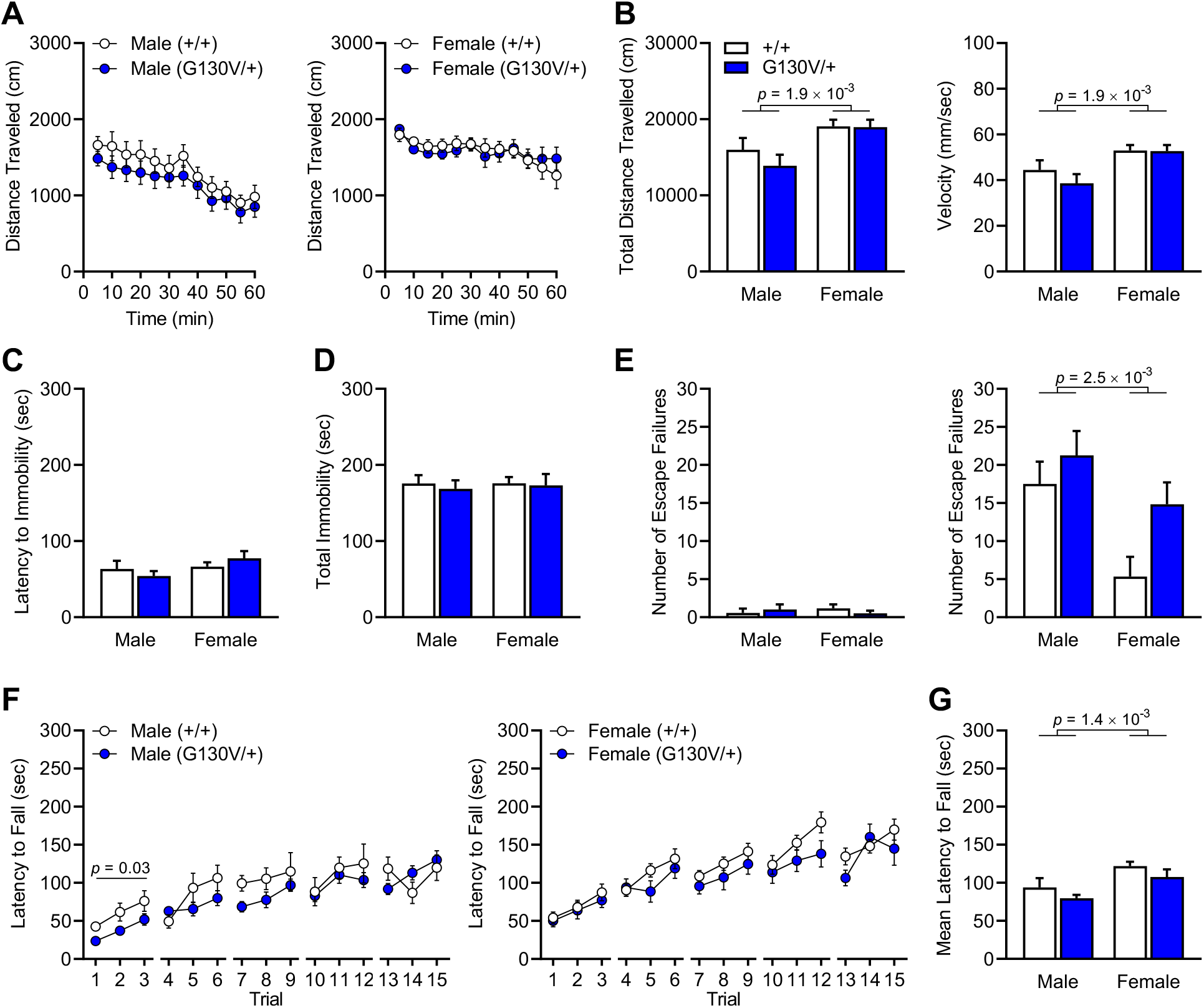
Behavioral characterization of *Dao*^*G130V/+*^ vs. *Dao*^*+/+*^ mice. A,B. Open field test: distance travelled per time, total distance travelled. C,D. Forced swim test: latency to immobility, total immobility. E. learned helplessness: left, number of escape failures in response to escapable shock in shock-naïve mice; right, number of escape failures in response to escapable shock in trained mice who were previously exposed to repeated inescapable shock. F,G. rotarod motor learning: latency to fall per trial, mean latency to fall.

A major risk factor for the onset of psychiatric disorders, including bipolar disorder, is exposure to stress *(48, 49)*. To test whether the *Dao*^*G130V*^ variant confers enhanced stress susceptibility, we assessed how well mice adapt to both forced swimming and foot shock stress. The forced swim test was used to assess escape-directed behavior in response to an acute inescapable swimming stress lasting six minutes. No differences in the latency to become immobile (Fig. 4C) were detected as a function of sex (*F*_(1, 43)_ = 2.4, *P* > 0.1) or genotype (*F*_(1, 43)_ = 0.01, *P* > 0.1). Similarly, no differences were detected in the total time spent immobile (Fig. 4D), as a function of either sex (*F*_(1, 43)_ = 0.04, *P* > 0.1) or genotype (*F*_(1, 43)_ = 0.18, *P* > 0.1). These data suggest that heterozygous *Dao*^*G130V*^ mutations do not enhance susceptibility to acute swimming stress. However, when mice were exposed to repeated inescapable foot shock in the learned helplessness test, there was a significant main effect of genotype (F_(1, 43)_ = 5.2, *P* = 2.7e-2) as well as sex (*F*_(1, 43)_ = 10.3, *P* = 2.5e-3) on the number of times mice failed to escape shock during testing when an escape route was provided (Fig. 4E, *right*). Specifically, *Dao*^*G130V/+*^ genotype and male sex were each associated with increased escape failures, with an insignificant sex × genotype interaction (*F*_(1, 43)_ = 0.98; *P* > 0.1). These data suggest that the *Dao*^*G130V*^ variant enhances susceptibility to repeated inescapable foot shock stress.

Additional control experiments refine our understanding of this effect. Importantly, the effects of sex and genotype on escape failures were not driven by differences in shock sensitivity, as shock-naïve mice of both genotypes were equally as likely to elude escapable shock (Fig. 4E, *left*). In addition, despite differences in stress susceptibility (Fig. 4E, *right*), all mice displayed a comparable hedonic response to natural reward in the sucrose preference test, regardless of sex (*F*_(1, 43)_ = 0.35, *P* > 0.1) or genotype (*F*_(1, 43)_ = 0.19, *P* > 0.1; Fig. S3C). This suggests that the ability to experience pleasure remains intact in *Dao*^*G130V/+*^ mice, even after repeated inescapable shock exposure (Fig. S3D). Similarly, no differences in nest construction in the nest building test were detected among the groups before or after the learned helplessness test (Fig. S3E-H). Taken together, these results suggest a specific effect of heterozygous *Dao*^*G130V/+*^ variants on susceptibility to some types of stress and not on other aspects of sensitivity to aversive or pleasurable stimuli.

Learned helplessness can be interpreted as an inability to learn the degree to which an adverse outcome is dependent on responses *(50, 51)*. To evaluate the ability of *Dao*^*G130V/+*^ mice to learn in a non-emotional context, we utilized the rotarod test (RRT), a cerebellum-dependent motor learning task. In males, there was a significant effect of time (*F* = 17.26, *P* < 1e-4) and a time × genotype interaction (*F*_(14, 266)_ = 1.9, *P* = 2.5e-2) on latency to fall across the five days of the procedure (Fig. 4F, *left*). When assessing comparisons within a given day of training, there was a main effect of genotype on the first day of training in male mice (*F*_(1, 19)_ = 6.0, *P* = 0.023), which suggests that the *Dao*^*G130V*^ variant led to an initial modest reduction in motor performance. In females, there was a significant main effect of time (*F* = 22.97, *P* < 1e-4) on the latency to fall across the five days of the procedure (Fig. 4F, *right*), which suggests that both males and females improved in their motor learning over the five days of training. Overall, there was a significant main effect of sex (*F*_(1, 41)_ = 11.7, *P* = 1.4e-3) and a trend effect of genotype (*F*_(1, 41)_ = 3.0, *P* = 0.09; Fig. 4G) on the latency to fall, which suggests that heterozygous *Dao*^*G130V*^ variants only modestly influenced motor learning in this task.

Since DAO^G131V^ had decreased enzymatic activity to degrade *D*-serine (Fig. 2E,F), a co-agonist of the *N*-methyl-D-aspartate glutamate receptor (NMDAR), we predicted that *Dao*^*G130V*^ may modulate NMDAR inhibition-dependent hyperlocomotor responses. To test this, we systemically administered MK-801, which is a non-competitive NMDAR antagonist that stimulates locomotor activity through a dopamine-independent mechanism *(52)*. Prior to MK-801 injection, baseline locomotor behavior did not vary as a function of genotype in male or female mice (*F*_(1, 21)_ = 0.08, *P* > 0.1; *F*_(1, 22)_ = 0.39, *P* > 0.1, respectively; Fig. 5A), but did vary as a function of time (*F =* 13.08, *P* < 1e-4; *F* = 2.75, *P* = 2.0e-2, respectively). After MK-801 injection, we found an attenuated hyperlocomotor response in male *Dao*^*G130V/+*^ vs. *Dao*^*+/+*^ mice (*P* = 4.8e-2; Fig. 5A, *left*). By contrast, the hyperlocomotor response was not significantly different in female *Dao*^*G130V/+*^ vs. *Dao*^*+/+*^ mice (*F*_(1, 22)_ = 0.08, *P* > 0.1; Fig. 5A, *right*). We conclude that heterozygous *Dao*^*G130V*^ diminished the stimulatory actions of NMDAR antagonism, especially in male mice.

**Figure 5.**
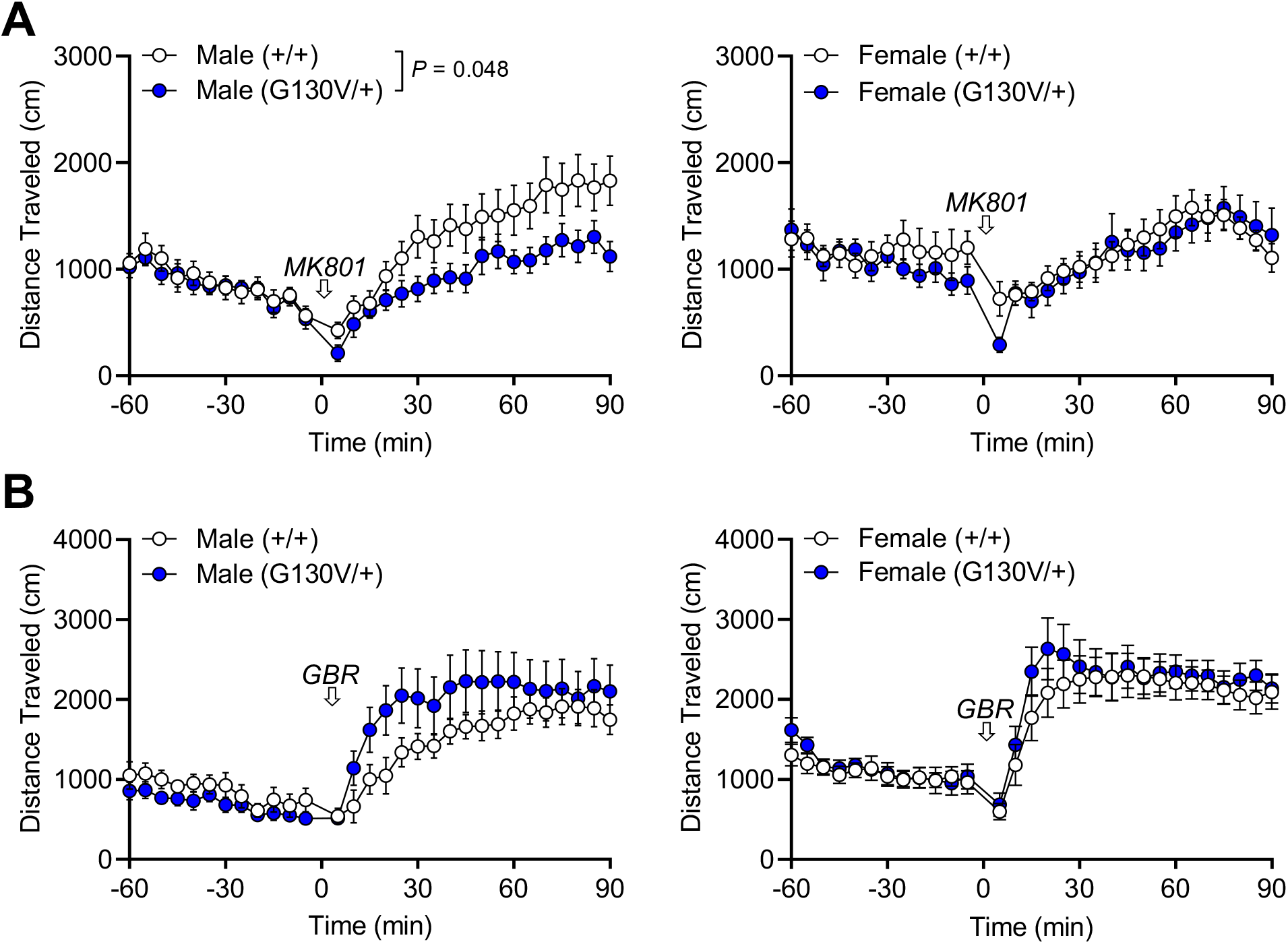
*Dao*^*G130V/+*^ blunts hyperlocomotor response to the NMDAR antagonist MK-801. A. Distance travelled per time before and after administration of the NMDAR inhibitor MK-801. B. Distance travelled per time before and after administration of the dopamine transporter blocker GBR12909.

To test the specificity of this effect, we assessed locomotor responses following administration of GBR12909, a dopamine transporter blocker that induces locomotor sensitization independent of the NMDAR. No significant effects of genotype were observed (*P* > 0.05, Fig. 5B). This result suggests that the attenuated hyperlocomotor response in male *Dao*^*G130V/+*^ mice does not involve dopamine and is more likely related to the effects of DAO on NMDAR transmission

### Associations of *Dao*^*G131V*^ and bipolar disorder with gene networks in the cerebellum

To gain insight into the molecular and cellular consequences of the *Dao*^*G131V*^ variant, we performed mRNA sequencing (RNA-seq) of cerebellar tissue from *Dao*^*G130V/G130V*^, *Dao*^*G130V/+*^, and *Dao*^*+/+*^ mice at 8 to 15 postnatal weeks. We tested for additive effects of *Dao*^*G131V*^ genotype and detected significant changes in the expression of 14 genes (FDR < 20%; Table S5), including *Ncam1*, encoding neural cell adhesion molecule 1, a positional candidate gene at risk loci from several recent GWAS of mood disorders *(53–55)* (*log*_*2*_ (fold change) = 0.38; *P* = 9.9e-5).

We integrated our RNA-seq data from *Dao*^*G130V*^ knock-in mice with previously published gene expression profiles of post-mortem cerebellar tissue from humans with major depressive disorder, bipolar disorder, schizophrenia, or no mental illness *(56)* to characterize shared patterns of transcriptional dysregulation. Using Weighted Gene Co-Expression Network Analysis (WGCNA) *(57)*, we identified 18 consensus gene co-expression modules with shared co-expression patterns in mouse and human cerebellum, involving a total of 6,107 genes (Table S5). We tested each module for changes in expression associated with *Dao*^*G130V*^ genotype and with psychiatric diagnosis (the three disorders combined). We then meta-analyzed the p-values to identify joint associations. Two modules were differentially expressed at an FDR < 0.05: M10 (199 genes, *P*_*meta*_ = 4.3e-4; Fig. 6A) and M5 (358 genes, *P*_*meta*_ = 2.6e-3, Fig. 6B). Both of these modules were down-regulated both in mice carrying the *Dao*^*G131V*^ variant and in psychiatric cases vs. controls (Fig. 6C-F).

**Figure 6.**
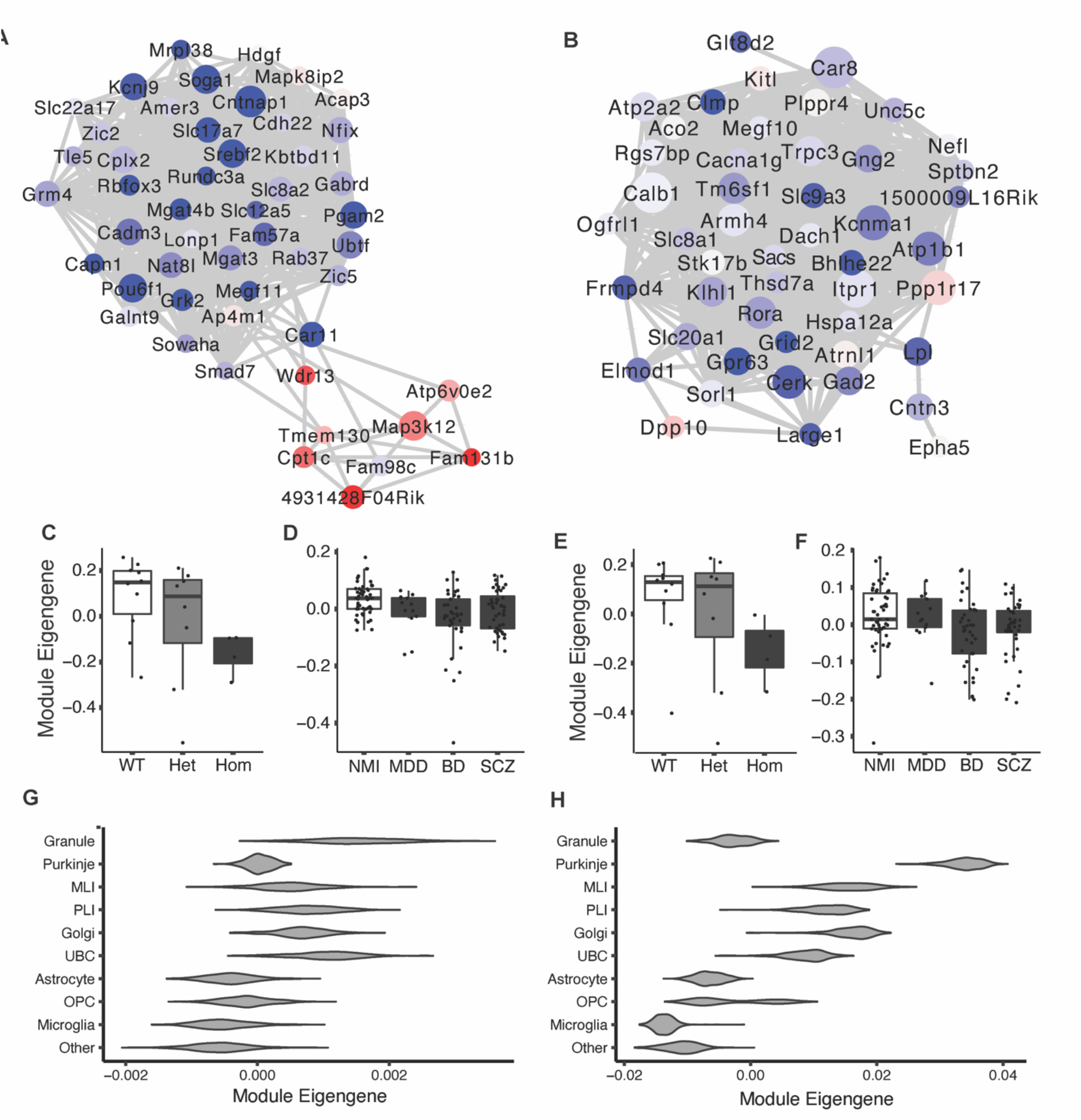
Differentially expressed gene networks in the cerebellum of *Dao*^*G130V/+*^ mice and of humans with bipolar disorder and schizophrenia. A,B. Module membership and differential expression of genes in modules M10 (A) and M5 (B). Node size corresponds to network centrality; node color corresponds to significance of differential expression (blue = down-regulated; red = up-regulated); edges correspond to Pearson correlations in mouse cerebellum. C-F. Expression of M10 (C,D) and M5 (E,F) module eigengenes in cerebellar tissue from *Dao*^*G130V*^ knock-in mice vs. wildtype controls (C,E) and in human psychiatric cases vs. unaffected controls (D,F). G,H. Expression of M10 (G) and M5 (H) module eigengenes in major cerebellar cell types. Abbreviations: WT, wildtype (*Dao*^*+/+*^); Het, heterozygous (*Dao*^*G130V/+*^); Hom, homozygous (*Dao*^*G130V/G130V*^); NMI, no mental illness; MDD, major depressive disorder; BD, bipolar disorder; SCZ, schizophrenia; MLI, molecular layer interneuron; PLI, Purkinje layer interneuron; UBC, unipolar brush cell; OPC, oligodendrocyte precursor cell.

Cell type specificity analysis using published scRNA-seq from mouse cerebellum *(44)* revealed that the M10 eigengene (its first principal component) was expressed most highly in granule neurons (Fig. 6G), and several of the most highly connected genes in the module (hub genes) were expressed almost exclusively in this cell type. Gene set enrichment analyses showed that M10 was enriched for synaptic genes (17 genes, 3.5-fold enriched, *P* = 2.6e-5; Table S6), including genes localized to both the pre-synaptic active zone (4 genes, 14.0-fold enriched, *P* = 2.8e-3) and the postsynaptic membrane (8 genes, 3.8-fold enriched, *P* = 5.3e-3). Granule neuron-specific synaptic genes in M10 with evidence of down-regulation *(P <* 0.05) in *Dao*^*G130V*^ knock-in mice and/or in mood and psychotic disorders include *Slc17a7*, encoding the vesicular glutamate transporter VGLUT1; *Grin2c*, encoding the GluN2C NMDA receptor subunit; *Gabrd*, encoding the delta subunit of the GABA_A_ receptor; and *Sema4c*, which encodes a semaphorin cell-cell signaling protein with roles in synaptic plasticity.

The M5 eigengene and many of the M5 hub genes were most highly expressed in Purkinje neurons (Fig. 6H). Like M10, M5 was enriched for synaptic genes (25 genes, 2.9-fold enriched, *P* = 6.7e-6; Table S6). The strongest enrichment was for components of the post-synaptic density (18 genes, 4.4-fold enriched, *P* = 7.9e-7) and genes localized to dendrites (27 genes, 3.2-fold enriched, *P* = 3.5e-7), but the module also included several genes specific to GABAergic pre-synapses. Purkinje neuron-specific synaptic genes in M5 with evidence of down-regulation in *Dao*^*G131V*^ knock-in mice and/or in mood and psychotic disorders included *Grid2*, which encodes the delta-2 ionotropic glutamate receptor, *Gad2*, which encodes glutamate decarboxylase 2, the enzyme responsible for GABA production, and *Slc32a1*, a vesicular inhibitory amino acid transporter. Taken together, our RNA-seq results reveal that *Dao*^*G130V*^ causes transcriptional down-regulation of synaptic genes expressed in granule neurons and Purkinje neurons, and these changes are recapitulated in the cerebellum of humans with mood and psychotic disorders.

To further explore the relationship of these gene networks with psychiatric disorders, we tested for overlap with known psychiatric risk genes and gene networks, as in our genomic sequence analyses, above (Table S6). Both M10 and M5 were enriched (FDR < 0.05) for genes that are also down-regulated in the prefrontal cortex of individuals with bipolar disorder, schizophrenia, and/or autism, suggesting shared molecular pathology across these brain regions. Both M10 and M5 were also enriched for targets of the neuronal RNA binding proteins FMRP, RBFOX1/2, RBFOX3, and CELF4, which likely reflects the functions of these RNA-binding proteins in transporting nascent RNAs to synapses. Finally, we found that M5 --but notably not M10 – was enriched for neuropsychiatric risk genes identified by exome and genome sequencing studies. These included developmental disorder genes compiled by the Deciphering Developmental Disorders consortium *(15)* (49 genes, odds ratio = 1.5, *P* = 4.9e-3), as well as genes associated with schizophrenia from SCHEMA *(17)* (25 genes with SCHEMA *P* < 0.05; odds ratio = 1.6, *P* = 0.03). Remarkably, the most significant overlap was with “Community 1” genes from our own bipolar disorder pedigrees, described above (17 genes, odds ratio = 3.1, *P* = 8.7e-5). This analysis highlights several variants that could potentially influence the function of Purkinje neurons. For instance, chr3:155838590 C/T is a novel missense variant in *KCNAB1* that was identified in all four sequenced bipolar disorder cases (and no unaffected individuals) from a four-generation pedigree. *KCNAB1* encodes a cytoplasmic subunit that modulates the biophysical characteristics of shaker-type voltage-gated potassium channels. In the cerebellum, *KCNAB1* is expressed almost exclusively in Purkinje neurons *(44)*. Overall, these results suggest that effects on Purkinje neuron synapses may be a recurring theme among variants contributing to risk for bipolar disorder.

## DISCUSSION

We found that hundreds of rare, potentially functional variants in genes with synaptic and nuclear functions co-segregate with bipolar disorder in 41 multiply affected pedigrees. Although we and others have described rare variants co-segregating with bipolar disorder in multiply affected families, very few of these variants have been characterized experimentally *(7, 11)*. Indeed, the very large number of variants and genes detected in these pedigrees (we report an average of ∼20 variants per pedigree) has left it unclear whether any of these variants is sufficient to cause mood phenotypes in isolation from the polygenic background. To address this, we characterized the top-scoring variant identified in our analysis, *DAO*^*G131V*^, in human cells and in a genetically precise knock-in mouse. Functional characterization of *DAO*^*G131V*^ indicated that it causes reduced DAO protein abundance and enzymatic activity, increased stress susceptibility, blunted behavioral responses to pharmacological inhibition of NMDAR, and transcriptional down-regulation of synaptic gene networks in the cerebellum. These findings have implications for understanding the genetics and neurobiology of bipolar disorder.

We found that *DAO*^*G131V*^ leads to hypofunction of the DAO protein, with reduced enzymatic activity in HEK293T cells and lower abundance in human cells and in the cerebellum, medulla, and kidney of *Dao*^*G130V*^ knock-in mice. Protein-folding simulations suggested that *DAO*^*G131V*^ may increase free energy of a loop domain at a distance from the enzymatic pocket, so hypofunction may be due to misfolding and decreased protein stability. Heterozygous *Dao*^*G130V/+*^ knock-in mice displayed increased escape failures following inescapable foot shock. The *Dao*^*G130V*^-dependent reduction in escape-directed behavior was specific to a more intense stress exposure, as both male and female heterozygous mice do not manifest deficits to acute inescapable swimming stress compared to wild-type controls. These results suggest that *Dao*^*G130V*^ may confer an enhanced susceptibility to more severe forms of stress. Severe forms of stress are a major risk factor for bipolar disorder *(49, 58)*. Thus, it is plausible that altered stress susceptibility could contribute to risk for bipolar disorder in human carriers of *Dao*^*G131V*^.

Not surprisingly, the behavioral changes in heterozygous *Dao*^*G130V/+*^ mice were more subtle and specific than published findings in mice with homozygous DAO loss-of-function variants. Unlike homozygous mice *(46, 47)*, heterozygous *Dao*^*G130V/+*^ mice manifested no significant differences in innate anxiety-like responses. *Dao*^*G130V/+*^ mice also showed normal locomotor behavior, novelty exploration, nest building, preference for natural reward, and cerebellar-dependent motor learning. These results suggest that heterozygous *Dao*^*G130V*^ does not influence basal behavioral responsivity to normal environmental stimuli or learning in non-emotional contexts. Since all known human carriers of *Dao*^*G131V*^ are heterozygous, our findings may more accurately model aspects of DAO function that are relevant specifically to bipolar disorder in humans. In addition, in the context of a multiply affected pedigree, *Dao*^*G130V*^ co-occurs with other common and rare functional variants, and it is likely that the combined effects of these variants are required to explain the high rate of bipolar disorder and other mood disorders in this pedigree. Nonetheless, our results demonstrate that even in isolation from other genetic and environmental factors, heterozygous *Dao*^*G130V*^ is sufficient to cause changes in behavior related to bipolar disorder endophenotypes *(59, 60)*.

Convergent evidence suggests that *Dao*^*G131V*^ causes dysregulation of NMDAR and related components of glutamatergic synapses in the hindbrain. At a molecular level, *Dao*^*G131V*^ impaired degradation of *D*-serine *in vitro*. It is likely that *Dao*^*G131V*^ also results in elevated levels of *D-*serine and NMDAR activity in the hindbrain, as has been observed previously in heterozygous and homozygous mice with other DAO loss-of-function mutations *(32, 34)*. Direct evidence for dysregulation of NMDAR signaling comes from blunted hyperlocomoter responses after pharmacological inhibition of NMDAR. This result extends previous findings in homozygous mice with *Dao* loss-of-function mutations *(31)*. At a transcriptional network level, we found down-regulation of gene networks in the cerebellum that were enriched for components of glutamatergic synapses, including the NMDAR subunit *Grin2c* (GluN2C). In the cerebellum, D-serine also binds and modulates a distinct ionotropic glutamate receptor, the δ2 ionotropic glutamate receptor (GluD2) to regulate synaptic plasticity *(61, 62)*. The gene encoding GluD2, *Grid2*, is a hub gene of a down-regulated gene network in *Dao*^*G130V*^ knock-in mice. We speculate that transcriptional down-regulation of these genes is a compensatory response to sustained, elevated levels of *D*-serine, but additional experiments will be needed to establish the precise changes in synaptic physiology.

Our transcriptomic data suggest that dysregulated gene networks are expressed primarily in granule neurons and Purkinje neurons. Axonal projections from granule neurons, termed parallel fibers, form synapses on the dendrites of Purkinje neurons, with each fiber synapsing onto hundreds of thousands of Purkinje neurons. Plasticity of parallel fiber synapses – in part mediated by *D*-serine, NMDAR, and GluD2 – is critical for cerebellar learning *(63)*. Emotional learning in the cerebellum, as well as social behavior and cognition, is mediated by specialized circuits in the cerebellar vermis *(64)*. In fear conditioning paradigms, repeated foot shock resulted in long-term potentiation of parallel fiber synapses in the vermis, and this effect was attenuated in GluD2 knockout mice *(65, 66)*. The aversive stimulus in fear conditioning is very similar to foot shock in the learned helplessness test. Therefore, it is plausible that the increased stress susceptibility of *Dao*^*G130V/+*^ involves a deficit in parallel fiber synaptic plasticity.

In recent years, several studies have described rare variants that co-segregate with bipolar disorder in large, multiply affected pedigrees, including our own previous report on these 41 pedigrees *(4–8)*. A general conclusion from these studies has been that multiply affected pedigrees contain functional variants in many genes, enriched in broad categories relevant to neuronal functions. Similar conclusions can be drawn from exome and genome sequencing studies of bipolar disorder case-control cohorts *(9, 10)*. The variants that we found in synaptic genes (Community 1 in our analysis) are consistent with these previous findings. Functional enrichments for nuclear functions such as DNA damage responses and chromatin remodeling (Community 2) are also consistent with sequencing studies *(16)* and GWAS *(26)* of psychiatric disorders. Notably, protein-truncating variants in many of these same genes are associated with autism and developmental disorders, whereas the vast majority of variants that co-segregated with bipolar disorder in our dataset are missense or non-coding. Therefore, bipolar disorder may be associated with less deleterious variants in some of the same genes that cause neurodevelopmental disorders.

Evidence supporting the involvement of the cerebellum in bipolar disorder and related psychiatric disorders spans >25 years of research *(67–69)*. Human brain imaging studies indicate structural and functional changes in bipolar disorder and schizophrenia, including reduced cerebellar volume *(70–72)* and altered cortico-cerebellar functional connectivity *(72–74)*. Anatomical studies have revealed that Purkinje neurons are reduced both in number and in size in patients with psychiatric disorders *(68, 75–77)*. Our transcriptomic analyses are also consistent with previous studies demonstrating down-regulation of transcripts and proteins for synaptic genes, including GABA_A_ receptors and NMDA receptor components *(78–80)*. Our results suggest that rare variants acting primarily through these hindbrain circuits may contribute substantially to risk for bipolar disorder.

## MATERIALS AND METHODS

### Study design

The purpose of this study was to identify and characterize rare variants contributing to risk for bipolar disorder. The starting point for our analysis were genome sequences from multiply affected pedigrees with bipolar disorder. The 41 pedigrees were selected from a larger collection of 972 pedigrees collected by the NIMH Genetics Initiative and by sites at the University of California, San Diego, the University of California, San Francisco, and the University of Chicago. We identified variants within these pedigrees using linear mixed model association tests *(5)* and fine-mapping of pedigree-specific linkage regions *(12)* and contextualized variants in a protein interaction network. The *DAO*^*G131V*^ variant was selected for functional validation because it had the best combined score derived by combining variant rankings from genetic linkage/association (best p-value from linear mixed models and linkage analysis), predicted deleteriousness (CADD score), and eigencentrality in the protein interaction network (details in Supplementary Materials and Methods). Heterologous expression of *DAO*^*G131V*^ HEK293T cells was used to characterize its effects on protein abundance and DAO enzymatic activity. We generated a genetically precise knock-in mouse model of *DAO*^*G131*^, *Dao*^*G130V/+*^, to study its effects *in vivo. Dao*^*G130V/+*^ mice were characterized to determine effects on *Dao* mRNA expression, DAO protein abundance, behavior, and transcriptional networks in the cerebellum.

### Genome sequences

We analyzed published genome sequences of 200 individuals from 41 multiply affected families with bipolar disorder *(5)* to identify functional rare or uncommon variants that co-segregate with bipolar disorder. Whole-genome sequencing was performed to >40x depth by Complete Genomics (San Jose, CA). Details of this cohort and sequencing strategy have been described previously *(5, 12)*.

### Validation of *DAO*^*G131V*^ genotypes in human genomes. *DAO*^*G131V*^

(rs768676371) genotypes in members of the multiply affected pedigree, initially determined by genome sequencing and imputation from linkage scans, were validated using custom TaqMan assays run on an ABI 7900HT Fast Real-Time PCR System (Thermo Fisher Scientific, Waltham, MA). Forward primer: ACCTTTATTTCCACCTTTTGCTTACTGT; reverse primer: CCCTCCAGAATTAGGCTTGTGT; Reporter 1 sequence: CTCTAGCTATGGCTGGTTC; Reporter 2 sequence: ACTCTAGCTATGTCTGGTTC; VIC and FAM were used as due for reporters 1 and 2, respectively. One member of the family, a nephew of the proband with a diagnosis of bipolar disorder type 1 shown at right in Fig. 1G, was newly ascertained as part of the current study. A blood sample was collected from this individual and used for genotyping. The remaining genotypes were determined using genomic DNA from lymphoblastoid cell lines obtained from the Coriell Institute for Medical Research, Camden, NJ.

### Protein structure-based modeling of the DAO G131V variant

The thermodynamic impact of the G131V single-nucleotide variant (SNV; chr12:109283989 G/T) in DAO was estimated using protein structure-based simulations of protein stability *(81)*. Modeling was performed using various DAO crystal structures of obtained from the Protein Data Bank *(82)*, including: 2DU8 *(83)*, 2E48 *(83)*, 2E49 *(83)*, 2E4A *(83)*, 2E82 *(83)*, 3CUK*(84)*, 3G3E *(85)*, 3W4I *(86)*, 3W4J *(86)*, 3W4K *(86)*, 3ZNN *(87)*, 3ZNO *(87)*, 3ZNP *(87)*, 3ZNQ *(87)*. For both wild-type (WT) and mutated proteins, the sidechain and backbone conformations of each protein domain were sampled at least one hundred times using a Monte Carlo procedure to minimize the estimated total free energy of folding for the protein *(88)*. The estimated change in energy upon mutation (ΔΔG) was calculated as the total free energy of the mutated protein minus that of the wild-type protein.

### Site-directed mutagenesis

The hDAO missense variant G131V was introduced into the human DAO cDNA (GenBank sequence: BC029057) by site-directed mutagenesis (SDM) and cloned into a pCMV3-C-Myc vector (HG13372-CM, Sino Biological) using the Quick Change II Site-Directed Mutagenesis kit as per manufacturer’s recommendations (Agilent Technologies). In brief, complementary primer pairs (Supplementary Table S1) were designed to anneal to DAO cDNA with one of the base pairs (g>t at position 392) being mismatched to the WT sequence, resulting in a glycine-to-valine amino acid substitution in the DAO protein sequence at position 131. PCR reactions were conducted in a PTC-200 Peltier Thermal Cycler (MJ Research), and the program consisted of one cycle of 95 °C for 1-min, twelve cycles of 95 °C for 30-sec, 55 °C for 1-min, 68 °C for 12-min and one further cycle of 68 °C for 7-min. Following temperature cycling, the reaction was placed on ice for 2-min and then treated with 1 µl DpnI restriction enzyme (10U/µl, Agilent Technologies) and incubated for 1-hr at 37 °C to digest parental supercoiled dsDNA. Approximately 8 µl DpnI digested PCR product was transformed into chemically competent E. coli strain DH5α. To confirm the presence of the desired mutation, four colonies were picked randomly and extracted plasmids were sequenced using primers listed in Supplementary Table S1. After confirmation, the plasmid was transfected into a HEK293T cell line for functional and biochemical characterization. The plasmids were also fully sequenced to confirm that there are no other mutations in the gene except for the desired one introduced by SDM.

### Cell culture and transient transfection

The HEK293T cell line (HCL4517, Dharmacon) were maintained to reach near confluence in flasks containing DMEM (10569010, ThermoFisher scientific) supplemented with 10% fetal bovine serum (F4135, Sigma-Aldrich) at 37 °C in 5% CO_2_ incubator. One day prior to transfection, cells were plated at a seeding density of 5 × 10^5^ cells to reach a confluency of 50-70% on the day of transfection. The cells were transfected with 7.5 µg of pCMV3-hDAO-C-Myc construct, encoding the WT or G131V missense variant of hDAO using Xfect transfection reagent (631317, Takara Clontech). The HEK293T cells were also transfected with 7.5 µg of pCMV3-C-Myc negative control vector (CV014, Sino Biological) as a negative control plasmid. The growth medium was replaced 4-hr post-transfection.

### Generation of a knock-in mouse model for the *DAO*^*G131V*^ variant

Cyagen (Santa Clara, CA) was contracted to generate a genetically precise knock-in mouse carrying the *DAO*^*G131V*^ variant at the endogenous mouse locus, using Cas9 genome editing. Exon5 of the mouse *DAO* gene located on mouse chromosome 5 was selected as target site for Cas9 genome editing. The G130V (GGC to GTC) mutation sites in donor oligo was introduced into exon 5 by homology-directed repair (Supplemantary Fig S2). Cas9 mRNA, guide RNA (gRNA) generated by *in vitro* transcription, and donor oligo were co-injected into fertilized eggs for KI mouse production (Supplemantary Table 1). The pups were genotyped by PCR, followed by sequence analysis and TaiI restriction analysis. We received the F1 heterozygous founders from Cyagen and expanded this colony for use in *in vivo* experiments. All experimental procedures using these mice were approved by the University of Maryland Baltimore Animal Care and Use Committee and were conducted in full accordance with the National Institutes of Health Guide for the Care and Use of Laboratory Animals.

### Genotyping of *Dao*^*G130V*^ knock-in mouse line

All pups were weaned on postnatal day (PD) 23, and tail clips were collected. Crude lysates were prepared by overnight incubation of the tail clips in direct lysis reagent (Viagen Biotech) and proteinase K (Viagen Biotech) at 55 °C. On the following day, the reaction was deactivated by incubation at 85 °C for 45-min. The PCR products were generated from PCR genotyping using the genotyping primers listed in Supplementary Table1. The amplicons were then purified by ExoSAP-IT PCR product clean up kit (Thermo Fisher) followed by Tail restriction analysis (Thermo Fisher).

### Determination of *DAO* mRNA levels in transfected cell line and *Dao*^*G131V*^ knock-in mice

Total RNA was isolated from transfected cell line and from four different mouse brain tissues using RNeasy Plus Mini Kit (74134, Qiagen) and Nucleospin RNA isolation kit (Clontech) as per manufacturer’s recommendations). 500 ng of total RNA was reverse transcribed (RT) using Maxima H Minus cDNA Synthesis Master Mix (M1681, Thermo Scientific). The RT product was then subjected to quantitative real time PCR (qRT-PCR) using KAPA SYBR FAST qPCR Master Mix Kit (KK4601, Kapa Biosystems), and 10 µM each of forward and reverse transcript-specific primers (Table S1) were used for the qRT-PCR. PCR reactions were conducted in CFX 384 real time system (Biorad) and the program consisted of one cycle of 95 °C for 3 min, 40 cycles of 95 °C for 3-sec, 60 °C for 40 sec (annealing and polymerization) followed by melting curve analysis. Data were analyzed using Bio-Rad CFX Maestro software (Bio-Rad). The expression patterns of ActB, GAPDH, and RPL13A was used to normalize the qRT-PCR data. The real-time quantification of target and housekeeping transcript accumulation was performed in separate reactions. The CT values obtained by RT-qPCR were used to calculate the accumulation of target gene (relative mRNA accumulation), relative to geometric mean of ActB, GAPDH, RPL13A transcript, by 2^-ΔΔCt method, where ΔΔCt = (Ct, Target gene - Ct, ACT1) *(89)*.

### Determination of DAO protein levels in transfected cell line and *Dao*^*G131V*^ knock-in mice

Mouse brain tissues *viz*. cerebellum, medulla, hippocampus, cortex, as well as kidney, were dissected, flash frozen in liquid nitrogen, and stored at -80 °C. Cell lysates and tissue lysates were prepared in ice cold radioimmunoprecipitation assay (RIPA) lysis buffer (Cat#, Company) containing Halt Protease and phosphatase inhibitor cocktail (78440, Thermo Scientific) and protein concentration was determined using Pierce BCA Protein Assay Kit (Thermo Scientific) per manufacturer’s recommendations. The cell (16 µg) and the tissue lysates (100 µg brain; 50 µg kidney) were then subjected to electrophoresis using Bolt Mini Gels (Life Technologies) and blotted on a polyvinylidene difluoride membrane (Company) using Mini Blot Module transfer system (Life technologies) as per manufacturer’s recommendations. The membranes were blocked with TBS blocking buffer (Licor) for 1-hr prior to overnight incubation with primary antibodies (Supplementary Table 2). On the following day, the membranes were washed four times with TBS-t (1X TBS containing 0.1% Tween 20) followed by 1-hr incubation with Licor IRDye secondary antibodies (Supplementary Table 2) at room temperature. The proteins were then visualized using Odyssey CLx Imaging system and the results analyzed with Image Studio software.

### DAO Activity Assay

DAO activity was assayed as described by *(90)*. Briefly, the transfected cells were harvested and homogenized in ice cold sodium phosphate buffer (50 mM Na_2_ HPO_4_ ; pH 7.4). Ten µl of the homogenate was added to sodium phosphate buffer containing 100 µM Amplex red, 0.25 U/ml horseradish peroxidase, 50 mM D-serine, with or without 10 µM FAD, to a final volume of 20 µl. After 1-hr incubation at 37 °C, the fluorescence was measured with Tecan Reader (Company) at an excitation and emission wavelength of 544 nm and 590 nm, respectively. Controls were run on each plate, including a control without cell homogenate, a control without D-serine, and a control with 100 µM of the DAO inhibitor, 6-methyl-benzo[d]isoxazole-3-ol (MolPort-027-640-512). The fluorescence values of cell homogenates were calculated by subtracting their fluorescence values from fluorescence values of controls without D-serine.

### Immunofluorescence

Immunostaining was performed as previously described *(91)*. Briefly, HEK293T cells were plated on coverslips at a seeding density of 2 × 10^5^ cells and transfected with 7.5 µg of pCMV3-hDAO-c-Myc construct encoding either WT or the G131V missense variant of hDAO. The HEK293T cells were also co-transfected with 7.5 µg of pCAG-SpCas9-GFP-U6-gRNA (Addgene plasmid #79144) to further confirm transfection efficiency by looking at GFP expression. The growth medium was replaced 4-hr post-transfection. The coverslips were subsequently washed with 1XPBS (Invitrogen) 48-hr post-transfection and fixed with 4% paraformaldehyde (Electron Microscopy Sciences) for 10-min at room temperature. Fixed cells were permeabilized using 0.1% TritonX 100 (Sigma) and unspecific sites blocked with 10% donkey serum for 1-hr at room temperature. The cells were then incubated overnight at 4 °C with mouse monoclonal anti-myc (1:1000 Abcam ab32) primary antibody, followed by incubation with secondary antibody donkey anti-mouse Alexa568 (Molecular Probes). Coverslips were imaged in Keyence BZ-X700 fluorescent microscope.

### Behavioral characterization of *Dao*^*G131V*^ knock-in mice

Male and female mice (8-12 weeks) were group housed under standard conditions (12-h light-dark cycle) with food and water available *ad libitum*. Testing was conducted in order of the least to most stressful procedure (Fig. S3I); *(92)*. All experiments were conducted, analyzed, and scored by experimenters blind to the conditions.

### Sucrose Preference Test

The sucrose preference test (SPT) was used to assess hedonic responses to natural reward both before (baseline) and after the learned helplessness test (see below). Mice were single housed in a cage fitted with two identical drinking bottles, containing either water or a 1% sucrose solution. Mice were allowed to drink freely for two consecutive days, and the sucrose-containing bottle was counterbalanced across animals and day. Measurements were taken between 5:00-7:00 PM, and data from the second day were analyzed. Sucrose preference was calculated as a percentage of sucrose intake relative to the total volume of sucrose and water that was consumed. A reduction in sucrose consumption relative to water indicates a reduction in sucrose preference, which is an index of anhedonia, or a lessened ability to experience pleasure *(93)*. Sucrose preference was performed twice.

### Nest Building Test

The nest building test (NBT) was used to assess nest construction, which is sensitive to genetic mutations and environmental conditions that are thought to underlie pathological disease states *(94)*. Approximately 1-hr before the dark phase, mice were transferred into individual testing cages that contained wood-chip bedding and 3 g of new square nestlet material. On the following day, nests were given a score from 1-5, ranging from an untouched nestlet (1) to a complete nest (5), and any remaining un-shredded nesting material was weighed *(95)*.

### Open Field Test

The open field test (OFT) was used to assess locomotor activity and anxiety-like behavior *(96)*. Mice were individually placed into an open field arena (49 × 49 × 49 cm; San Diego Instruments, San Diego, CA) for 60-min (∼300 lux). The test was recorded by an overhead digital camera, and distance traveled, velocity, and time spent in the center area (as defined as the 50% most center area of the chamber) of the arena scored using TopScan (CleverSys Inc; Reston, VA). More time spent near the perimeter of the arena (i.e., thigmotaxis) as opposed to the center is considered an anxiety-like response in this task.

### Elevated Plus Maze

The elevated plus maze (EPM) was used to assess anxiety-like behavior, and is based on the innate desire of mice to be in enclosed spaces *(97)*. The EPM is comprised of two open arms and two closed arms (39 × 5 cm; Stoelting Co, Wood Dale, IL) elevated 50 cm above the ground. Mice were placed in the center zone of the area facing an open arm and allowed to freely explore the apparatus for 5-min (∼15 lux). The test was recorded by an overhead digital camera, and time spent in the open and closed arms was scored using TopScan (CleverSys Inc; Reston VA). Less time spent in the open arms is considered an anxiety-like response in this task.

### Forced Swim Test

The forced swim test (FST) was used to assess behavioral despair in response to an acute inescapable swimming stress *(98, 99)*. Mice were placed into a cylindrical transparent plexiglass tank (30 cm × 20 cm) filled with 15 cm of water (23 ± 1 °C). Mice were then recorded for 6-min by a tripod video camera. Latency to immobility and total immobility were manually scored, with immobility being the minimum movement that is required for the mice to keep their head above water. Mice typically engage in escape-directed behavior when placed into water but will eventually adopt a posture of immobility. Behavioral despair is indicated by a faster latency to immobility and an increased duration of immobility.

### Learned Helplessness Test

The learned helplessness test (LHT) was used to assess behavioral despair in response to repeated inescapable shock stress *(93)*. Coulbourn shuttle boxes were used, which contained two compartments separated by a guillotine door (Coulbourn Instruments LLC, Whitehall, PA). On the first day (*training*), mice were placed on one side of the box, and received 120 inescapable foot shocks delivered through an electric grid floor (0.3 mA, 2-sec shock duration, 15-sec intertrial interval). On the second day (*testing*), mice were placed on one side of the box, and received 30 escapable shocks that coincided with the guillotine door opening (0.3 mA, 15-sec shock duration, 20-sec average inter-trial interval). In the first five trials, the guillotine door opened once the shock was initiated, and remained open for the duration of the shock. In the remaining trials, the guillotine door opened with a 3 sec delay after initiation of the shock. In all trials, the shock was terminated as soon as the mouse escaped into the neighboring compartment. The number of escape failures were recorded and scored with Graphic State (Coulbourn Instruments, Whitehall, PA). To rule out an effect of genotype on shock sensitivity, naïve mice went through a similar procedure but were allowed to escape shock (Fig. 4E, *left*).

### Rotarod Test

The rotarod test (RRT) was used to assess motor learning and performance *(100)*. Mice were placed onto a rotarod beam (IITC Life Science Inc; Woodland Hills, CA) and were allowed to acclimate standing on the beam for one minute. The beam then began to rotate at 4 RPM, and progressively increased to 40 RPM by the end of the trial at 5-min. Three consecutive trials occurred on each day (2-min intertrial interval) for five consecutive days. Mice were recorded by a tripod video camera, and the latency to fall from the beam was scored for each trial on each day.

### MK-801-Induced Hyperlocomotion

MK-801 is a non-competitive *N*-methyl-D-aspartate receptor (NMDAR) antagonist that stimulates locomotor activity through a dopamine-independent mechanism *(52)*. Mice were habituated to an open field arena for 60-min (*see OFT above*). Mice then received a systemic injection of MK-801 (0.3 mg/kg, intraperitoneal, i.p.; Sigma-Aldrich), and were then returned to the open field for an additional 90-min. This dose and route of administration is what we have previously established to induce a moderate hyperlocomotor response in the absence of stereotypy *(101)*. MK-801 was dissolved in 0.9% NaCl on the day of testing. The test was recorded by an overhead digital camera, and distance traveled was scored using TopScan (CleverSys Inc; Reston VA).

### GBR12909-Induced Hyperlocomotion

GBR12909 is a dopamine transporter blocker that induces locomotor sensitization, independently of the NMDAR. Mice were habituated to an open field arena for 60-min (*see OFT above*). Mice then received a systemic injection of GBR12909 (16 mg/kg, i.p.; Sigma-Aldrich), and were then returned to the open field for an additional 90-min. This dose and route of administration is what we have previously established to induce a moderate hyperlocomotor response in the absence of stereotypy*(101)*. GBR12909 was dissolved in 10% dimethylsulfoxide (DMSO; Sigma-Aldrich) on the day of testing. The test was recorded by an overhead digital camera, and distance traveled scored using TopScan (CleverSys Inc; Reston VA).

### RNA sequencing

RNA-seq was generated from the cerebellum of 8- to 15-week-old *Dao*^*G130V/G130V*^ (n=2 male, 2 female), *Dao*^*G130V/+*^ (n=4 male, 4 female), and littermate C57BL/6N controls (n=6 male, 5 female). Total RNA was extracted from flash-frozen cerebellar tissue using the RNeasy Plus Mini kit (Qiagen). Strand-specific, dual unique indexed libraries for sequencing on Illumina Novaseq were made using the NEBNext® Ultra II™ RNA Library Prep Kit for Illumina®(New England Biolabs,Ipswich, MA). Manufacturer protocol was modified by diluting adapter 1:30 and using 3 ul of diluted adapter. The size selection of the library was performed with AMPure SPRI-select beads (Beckman Coulter Genomics, Danvers, MA). Glycosylase digestion of adapter and 2nd strand was done in the same reaction as the final library amplification. Sample input for this method was PolyA enriched RNA. Enrichment was done using the NEB Poly(A) mRNA magnetic isolation module. No changes were made to the manufacturer’s protocol. Libraries were sequenced to a depth of ∼50-75 million 100 bp paired-end reads on an Illumina NovaSeq6000 sequencer.

### Statistical analysis

All experiments were performed in a randomized fashion and conducted and analyzed by experimenters who were blind to the conditions. Biochemical and behavioral data were analyzed with GraphPad Prism Software 9.0.1. Distributions were assessed for normality using the D’Agostino-Pearson test and for homogeneity of variance using the Brown-Forsythe Test. Data are presented as mean ± standard error of the mean (SEM) and statistical significance was defined as *p* < 0.05. One-way analysis of variance (ANOVA) was used when data from three or more groups were being compared. Two-way ANOVA was used when sex (male vs. female) and *Dao*^*G130V/*^ genotype were independent factors. Two-way repeated measures ANOVA was used when genotype and time (5-min time bins; repeated measure) were independent factors. If a significant main effect and/or interaction was detected, the Holm-Šídák post-hoc test was used to assess pairwise comparisons. Methods for genome sequence analysis and RNA-seq data are provided as Supplementary Materials and Methods.

## Data Availability

Genome sequences have been deposited in the BIGPOWER database and are available upon request to Dr. Jared Roach. RNA-seq data have been deposited in the Gene Expression Omnibus (GSE# in progress). The DaoG130V/+ mouse line has been made available through the Mutant Mouse Resource and Research Centers (RRID:MMRRC_067164-UCD). All other data are available in the main text or the supplementary materials.

https://www.ncbi.nlm.nih.gov/gds

## Supplementary Materials

### Materials and Methods

Table S1. Annotation of variants co-segregating with bipolar disorder in multiply affected pedigrees

Table S2. Enrichment of genes from bipolar disorder pedigrees in neuropsychiatry gene lists

Table S3. Clustering and ranking of variants with protein interaction communities

Table S4. Enrichment of gene communities from bipolar disorder pedigrees for Gene Ontology terms

Table S5. Effects of *Dao*^*G130V*^ and psychiatric diagnosis on cerebellar gene expression: differential expression statistics and gene co-expression network membership

Table S6. Gene set enrichment analyses for consensus gene co-expression network modules M5 and M10

## Acknowledgments

We thank the Coriell Institute for providing DNA samples. We thank the staff of the University of Maryland Veterinary Resources and at the Mutant Mouse Resource and Research Center at UC Davis for assistance in maintaining and archiving the *Dao*^*G130V/+*^ knock-in mouse line. We thank the Genomics Resource Center at the University of Maryland School of Medicine for RNA sequencing. Most importantly, we thank the families who have participated in and contributed to these studies.

## Funding

National Institute of Mental Health grant R01 MH094483 (JRK)

National Institute of Mental Health grant R01 MH110437 (PPZ)

National Institute of General Medical Sciences Center for Systems Biology (P50 GM076547, Leroy Hood, principal investigator)

Intramural Research Program of the National Institute of Mental Health (FJM)

U.S. Department of Veterans Affairs Merit Awards I01BX004062 and 101BX003631 (TDG)

University of Luxembourg–Institute for Systems Biology Strategic Partnership (Leroy Hood, principal investigator)

A NARSAD Young Investigator Award from the Brain and Behavior Research Foundation (SAA)

Seed funding from the University of Maryland School of Medicine (SAA)

Data and biomaterials were collected as part of 11 projects (Study 40) that participated in the NIMH Bipolar Disorder Genetics Initiative (MH59545, MH059534, MH59533, MH59553, MH60068, MH059548, MH59535, MH59567, MH059556, and 1Z01MH002810-01), which was also supported by NIH Grants P50CA89392, from the National Cancer Institute, and 5K02DA021237, from the National Institute of Drug Abuse.

## Author contributions

Conceptualization: NH, SAA

Investigation: NH, LMR, TS, JA, RL, RTO, EMH, JB, PT, JCR, SAA

Funding acquisition: SAA, JRK, PPZ

Supervision: DCG, DWC, HWE, ESG, FJM, JIN, PPZ, JRK, JCR, TDG, SAA

Writing – original draft: NH, LR, SAA Writing – review & editing: all authors

## Data and materials availability

Genome sequences have been deposited in the BIGPOWER database and are available upon request to Dr. Jared Roach. RNA-seq data have been deposited in the Gene Expression Omnibus (GSE# in progress). The *Dao*^*G130V/+*^ mouse line has been made available through the Mutant Mouse Resource and Research Centers (RRID:MMRRC_067164-UCD). All other data are available in the main text or the supplementary materials.

## SUPPLEMENTARY MATERIALS

### Materials and Methods

#### Genome sequence analysis

Given our relatively modest sample size, our goal is not necessarily to identify associations that reach standard thresholds for genome-wide significance. Rather, we wish merely to identify the most plausible moderate-to large-effect variants within each pedigree. It is inevitable that some of the individual variants identified by these approaches will be false positives. Therefore, we have focused on identifying and replicating patterns at the level of gene sets, as well as functional validation. Variant analysis was performed in two phases. Initial variant annotation and statistic genetic analyses were performed in 2014 using annotations available at that time. The *DAO G131V* variant was initially identified and selected for experimental validation based on this first phase of analysis. Variant annotation resources have matured considerably since those initial analyses. We therefore updated our analysis in April, 2021 using current gene models, allele frequency databases, and variant deleteriousness scores, resulting in the re-annotation or elimination of a small number of variants. Network modeling and gene set enrichment analyses presented here were performed using the updated annotations.

The first phase of variant annotation was performed using QIAGEN’s Ingenuity Variant Analysis, Kaviar *(102)*, and custom variant annotation tools in the Family Genomics Toolkit (familygenomics.systemsbiology.net/software), as previously described *(5)*. The updated annotation was performed using the Ensembl Variant Effect Predictor, GRCh37 release 103 *(103)*. Because indels called from Complete Genomics genome sequences are not always reliable, our analysis focused on single-nucleotide variants. We selected variants with global allele frequencies < 5% in both 1000 Genomes Phase 3 *(104)*. We further selected variants with the following effects relative to both UCSC knownGene models (phase 1) and Ensembl release 103 protein-coding transcripts (update): stop gained, stop lost, start lost, splice donor variant, splice acceptor variant, missense, 3’ untranslated region variant, 5’ untranslated region variant. These variants were further annotated with CADD to predict their deleteriousness *(27)*.

Affection status models are described in our previous publication *(5)*. Briefly, we considered all individuals with a diagnosis of bipolar disorder type 1, bipolar disorder type 2, or schizoaffective disorder bipolar type to be affected. Individuals with recurrent or single episodes of major depression were also considered to be affected if (i) the best-supported pre-determined linkage peak for that pedigree *(12)* supported an inheritance model that included major depression, and they were not married-in; or (ii) they had offspring with BD and were not married-in to a pedigree.

Genetic associations of variants with bipolar disorder were calculated by two approaches. For variants with allele frequencies >1% in our dataset – primarily those in very large families or present in more than one pedigree – we computed associations with linear mixed models implemented with EMMAX *(105)*, controlling for the polygenic background. To improve statistical power, this analysis integrated the 200 genomes from bipolar disorder pedigrees with an additional 254 Complete Genomics genomes from individuals with no known psychiatric or neurological condition. This analysis extends an analysis of candidate genes described in our previous publication to all protein-coding genes, and additional details are described therein *(5)*.

Our study was designed to incorporate data from pre-determined linkage scans, and in many pedigrees we had genome sequences from only one or a few of several affected individuals. Thus, certain variants were identified in fewer than 1% of the genomes and therefore could not be included in the linear mixed model, despite the fact that they perfectly co-segregated with bipolar disorder in one of the pedigrees. We prioritized these variants by fine-mapping pre-determined linkage regions using the Family Genomics Toolkit, using pedigree-specific linkage peaks derived from linkage scans with 4,500 SNPs genotyped in a larger number of individuals from each pedigree *(12)*. This Toolkit is primarily designed to detect candidate variants in single pedigrees. For this analysis, we considered only coding variants, as we cannot rely as extensively on genetic association in variant selection.

Raw scores for each variant were based on genotype call quality score, allele frequency in the population, and functional predictions for the effect of an allele are combined into a single raw score. For each gene, the distribution of the best (i.e., lowest) variant score within that gene was tabulated across all best variants within genes observed transmitted from each parent to child in a control set of 484 trios from non-bipolar disorder pedigrees. A score was assigned to that gene as the weight of the single tail of that distribution equal to or more extreme than the observed score. Due to a pseudocount, the lowest employed score is the reciprocal of the number of genes, or approximately 1:25,000. Due to the large number of olfactory receptor genes and the lack of a general model for false positivity encompassing them, variants within olfactory receptors were assigned an impossibly high pseudocount.

Separately, we estimate the likelihood of a variant falling within an inheritance state consistent with segregation with a specified affectation status model. Here, since our goal is to fine-map pre-determined linkage regions, we also estimate this likelihood from the LOD score, *l*, of the closest previously genotyped SNP to that variant. This probability was computed as 10^−*l*^. The lower of this segregation probability or the segregation probability computed from WGS segregation was then employed for further analysis. Our workflow allows variants without perfect segregation to have a non-zero probability. However, in the current analysis only perfectly segregating variants are reported, as no other variants had sufficient overall score to reach a reporting threshold.

A combined probability for each variant was then computed by multiplying the annotation probability by the segregation probability. These two probabilities are independent. They are expected to be accurate representations of probability most of the time. Our informal application of Bayesian logic is designed to capture much information from multiple underlying datasets and analyses to compute a single statistic to enable relative ranking of candidate variants. Up to 10 variants (retaining variants tied for a particular overall probability) were reported for each pedigree. We set a strict threshold for overall variant probability of 2% located at a predetermined linkage region with a maximum LOD score >0.6, corresponding to a p-value ∼ 0.05. For five pedigrees no candidate variants were reported.

741 genes contained at least one variant with a p-value < 0.05 from the linkage and association analyses described above. We characterized these genes through genes set enrichment analysis and integration with protein-protein interaction networks. First, using Fisher’s exact tests, we calculated over-representation of these genes among 22 gene sets from genetic and genomic studies of neuropsychiatric disorders. The gene sets were as follows: (1) genes intolerant of loss-of-function variants from gnomAD *(13)*; (2-5) risk genes from studies of rare variants in four disorders, including severe developmental disorder risk genes from the Deciphering Developmental Disorders consortium’s DDG2P database *(15)*, autism spectrum disorder risk genes from the Autism Sequencing Consortium *(16)*, schizophrenia risk genes from the SCHEMA consortium *(17)*, and bipolar disorder risk genes from the BipEx consortium *(10)*; (6-9) genes identified from large-scale GWAS, identified by gene-based analyses with MAGMA *(106) (P <* 2.77e-6) for bipolar disorder *(2)*, major depression *(18)*, and neuroticism *(20)*, and epigenomic fine-mapping of risk loci for schizophrenia *(19)* (http://resource.psychencode.org/Datasets/Integrative/INT-18_SCZ_Risk_Gene_List.csv); (10-15) differentially expressed genes in the prefrontal cortex of individuals with schizophrenia, bipolar disorder, and autism from the PsychENCODE consortium *(21)* (http://resource.psychencode.org/Datasets/Derived/DEXgenes_CoExp/DER-13_Disorder_DEX_Genes.csv); (16-21) gene networks derived from functional genomics experiments to define targets of the neuropsychiatric risk genes FMRP, RBFOX1/3, RBFOX2, CHD8, CELF4, and microRNA-137, annotated by *(22)*; (22) synaptic genes from SynaptomeDB *(107)*. Table S2 provides gene membership for each of these sets.

Protein-protein interaction (PPI) networks were derived using data from GeneMANIA *(23)*. We downloaded the GeneMANIA combined human PPI network, which consists of 6,979,631 PPI interactions among human proteins from integration of a wide variety of underlying studies. Using these data, we first tested whether genes identified in bipolar disorder pedigrees are more likely to interact with each other in this network than expected by chance. We calculated two metrics to summarize these interactions: the total number of direct interactions among the 741 genes, and the sum of the interaction weights from the GeneMANIA database. We calculated an empirical p-value based on a background distribution derived by permuting the edges in the full GeneMANIA network 1,000 times. The empirical p-value for the observed number of interactions and edge weight sum is the number of permutations in which these metrics are larger than the observed metrics, divided by the total number of permutations. To avoid a p-value of zero, a pseudocount of 1 is added to both the numerator and the denominator.

Next, we searched for gene “communities” (modules) amongst our 741 genes. A community is defined as a group of genes with a high number of within-community PPI and a lower number of between-community PPI. The starting point for this analysis was a weighted graph of PPI from GeneMANIA, including direct interactions among the 741 genes, as well as “one-hop” interactions in which two genes from our list are connected via a shared interaction with any other human gene. In our primary analysis, direct interactions were assigned a weight 10x greater than one-hop interactions. We detected communities in this graph using Leiden clustering, with a resolution of 0.8, implemented using the igraph and leiden R packages *(24, 108)*. This analysis revealed two major communities, as well as two smaller communities with fewer than 20 genes, which were dropped from downstream analyses. We annotated each of the major communities using Gene Ontology terms derived from the org.Hs.egGO2ALLEGS object in the org.Hs.eg.db R package. Gene set enrichment analysis was performed using Fisher’s exact tests. In addition, we computed the centrality of each gene within its community based on eigencentrality, computed within igraph. Community detection was highly robust to the edge weight parameter. As expected, smaller sub-communities could be identified at higher resolution. Similar communities were also obtained using Louvain clustering. All of the major findings hold true in all of these versions of the analysis.

We developed a combined ranking system to prioritize variants for functional validation. First, we ranked variants by the p-value from linear mixed modeling or fine-mapping of linkage peaks (for the latter analysis, we derived this p-value from the best LOD score within the linkage region). Second, we ranked variants by their predicted deleteriousness, based on CADD scores *(27)*. Third, we ranked variants by the gene’s eigencentrality in the PPI network. The final rank is the unweighted average of these three scores.

**RNA-seq analysis. Following standard base-calling and adaptor trimming, we** used kallisto *(109)* for pseudo-alignment of sequencing reads to the mouse genome (mm10), estimation of read counts and transcripts per million in each transcript. Transcripts with a median TPM > 1 were retained for analysis. Principal component analysis and sample hierarchical clustering were used to detect outliers, and a single C57BL/6N male was removed. We used the glmFit() function in the edgeR R package to fit counts to a generalized linear model, including trended dispersion estimates *(110)*. We then performed likelihood ratio tests with the glmLRT() function to estimate additive effects of the *Dao*^*G130V*^ allele (0, 1, or 2 copies), including main effects of age and sex as covariates.

We compared the RNA-seq data from mouse cerebellum to published gene expression profiles of cerebellum from bipolar disorder, schizophrenia, and major depression cases vs. non-diseased controls *(56)*. The human gene expression profiles were produced using the Affymetrix Human Gene 1.0 ST Array transcript (gene) version. We downloaded normalized data from the Gene Expression Omnibus (GSE35974) and used probe annotations from the hugene10sttranscriptcluster.db R package. We used the lmFit() and eBayes() functions from the limma R package *(111)* to fit a linear model and estimate effects of diagnosis (any psychiatric diagnosis vs. non-diseased controls), including age, sex, post-mortem interval, and pH as covariates.

We reconstructed a consensus gene co-expression network to integrate our mRNA-seq data from *Dao*^*G130V*^ knock-in mice. We identified one-to-one orthologs in human vs. mouse using annotations from biomaRt, and filtered both datasets to a common set of genes that were detected in both datasets. Network reconstruction was performed using the blockwiseConsensusModules() function from the WGCNA R package *(57)*, with power = 6, corType = ‘bicor’, networkType = ‘signed’, minModuleSize = 30, mergeCutHeight = 0.3, and minKMEtoStay = 0.

Associations of modules with mouse genotype and with human psychiatric disease were computed with ROAST gene set tests *(112)*. For the mouse dataset, we fit a model to estimate additive effects of the *Dao*^*G130V*^ allele (0, 1, or 2 copies), including age and sex as covariates. For the human dataset, we fit a model to estimate effects of diagnosis (any psychiatric diagnosis vs. non-diseased controls), including age, sex, post-mortem interval, and pH as covariates. We computed a combined meta-analytic p-value for each module using Fisher’s method to combine p-values, and we selected modules with a significant meta-analytic p-value at a False Discovery Rate < 0.05 and the same prevailing direction of effect (down- or up-regulated) in both datasets. For visualization purposes, we also calculated the module eigengene (first principal component) in each dataset, using the moduleEigengenes() function from WGCNA.

Gene Ontology gene set enrichment analysis of modules M5 and M10 was performed with DAVID *(113)*. Gene set enrichment analyses with neuropsychiatric gene lists was performed using Fisher’s exact tests. Hub genes within each module were detected based on eigen centrality, computed in the igraph R package *(108)*.

To characterize the cell type-specific expression patterns of modules M5 and M10, we used single-nucleus RNA-seq data from mouse cerebellum *(114)*. Fully analyzed snRNA-seq data were downloaded from the Single Cell Portal (https://singlecell.broadinstitute.org/single_cell/study/SCP795/a-transcriptomic-atlas-of-the-mouse-cerebellum). We calculated the module eigengene expression of modules M5 and M10 within the snRNA-seq data. We then tested for enrichment of eigengene expression in each major cell type using the FindMarkers() function from the Seurat R package *(115)*.

**Figure S1.**
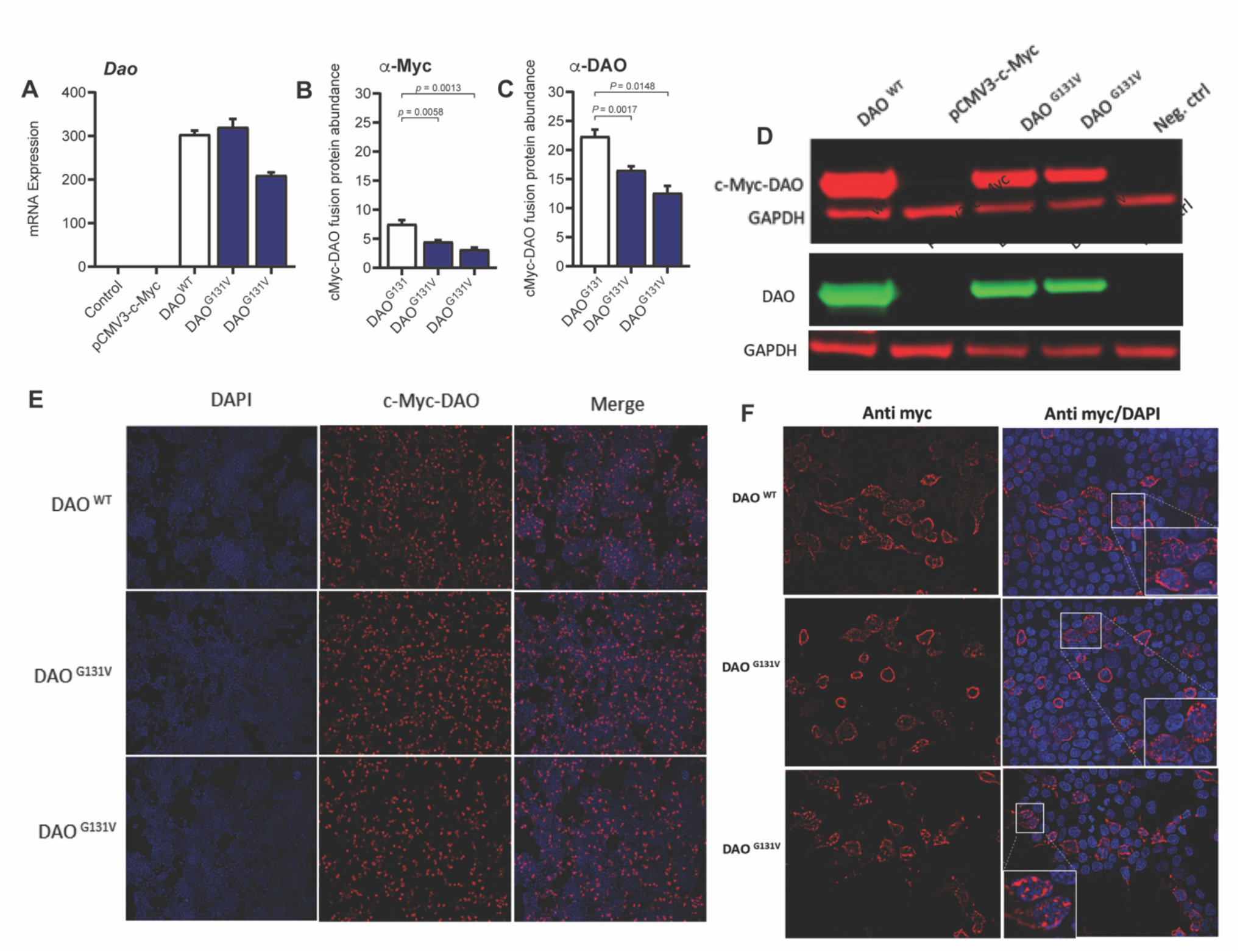
Ectopic expression of *DAO*^*G131*^ vs. *DAO*^*G131V*^ in HEK293T cells. c-Myc-DAO fusion protein with reference (G131) or variant (G131V) allele at position 131 was transiently expressed in HEK293T cells. A. mRNA expression. B. *Dao* mRNA expression (qPCR). B. DAO protein abundance quantified with an α-Myc antibody (Western blot). C. DAO protein abundance quantified with an α-DAO antibody (Western blot). Note: Quantitation in B and C is relative to GAPDH, whereas the plot in Fig. 2D is normalized to *DAO* mRNA levels to account for transfection differences. All quantitation approaches show a significant decrease in DAO protein. D. Blots from B (top) and C (bottom). E,F. DAO protein localization quantified by immunofluorescence.

**Figure S2.**
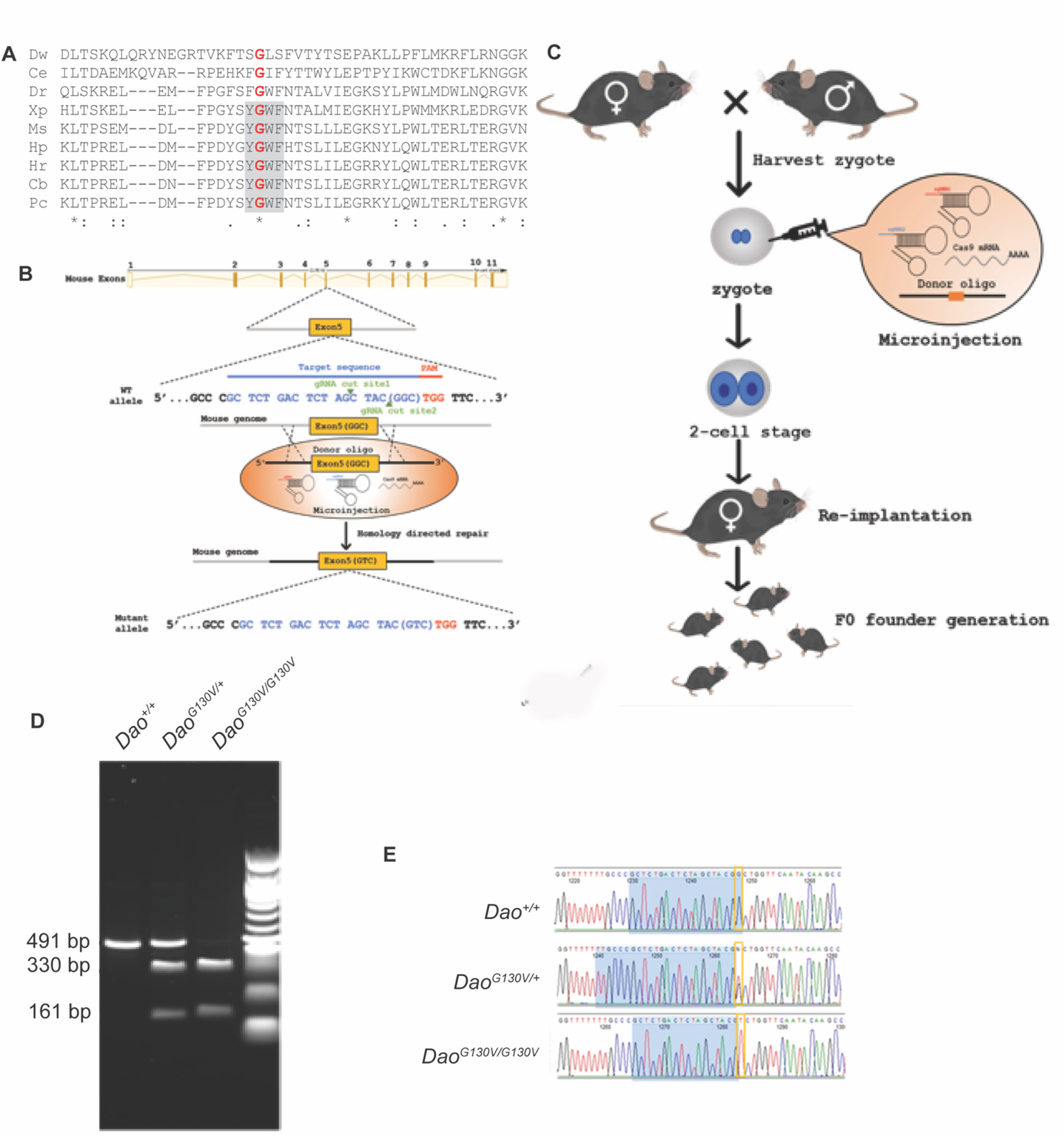
Generation of the *Dao*^*G130V/+*^ knock-in mouse model of the *DAO*^*G131V/+*^ variant. A. Evolutionary conservation of the *DAO*^*G131*^ residue. B. Genome-editing strategy. C. Breeding strategy. D,E. Genotyping of *Dao*^*G130V*^ by restriction digest (D) and Sanger sequencing (E).

**Figure S3.**
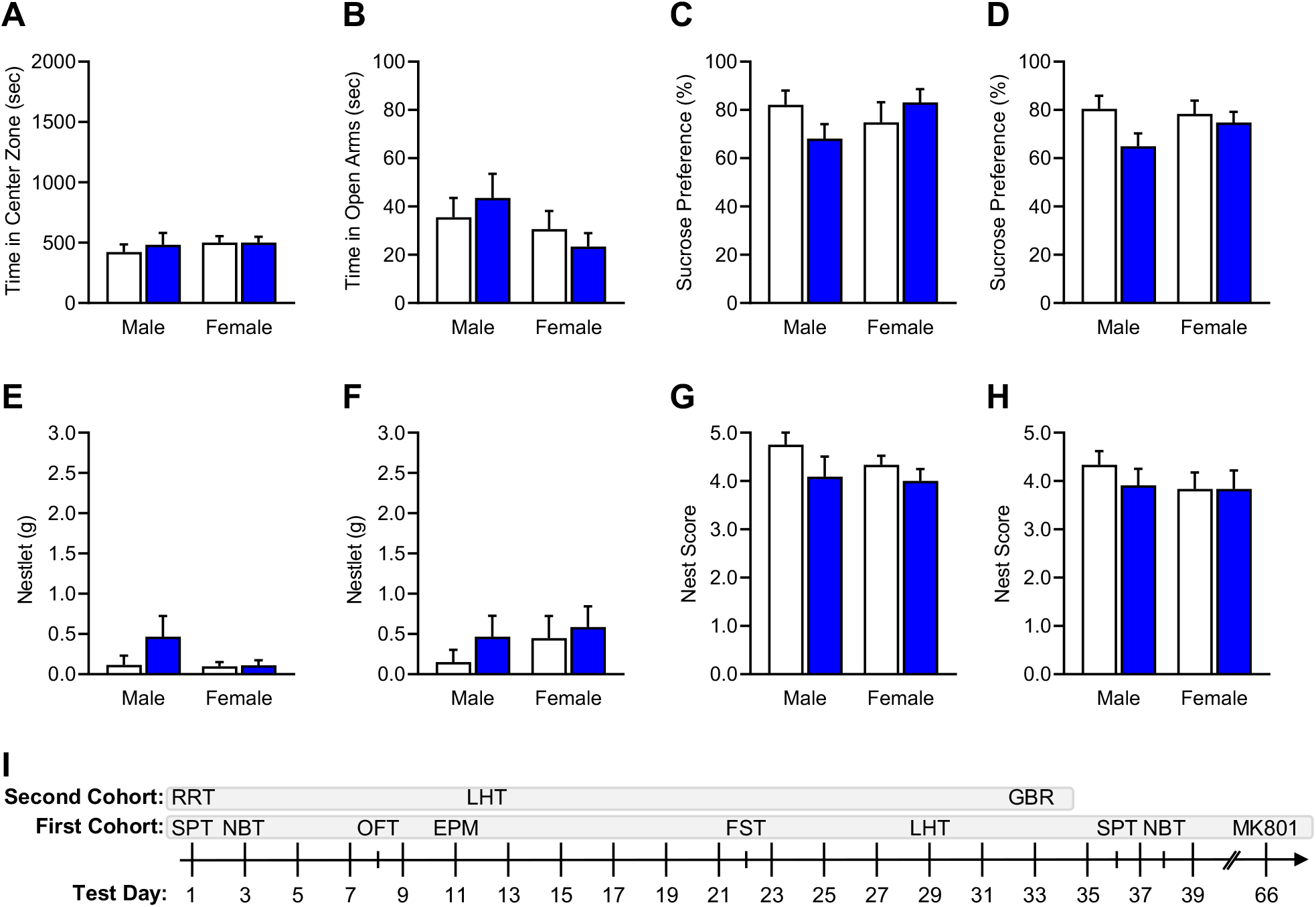
Behavioral characterization of *Dao*^*G130V/+*^ vs. *Dao*^*+/+*^ mice. **A.** Open field test: time spent in center. B. Elevated plus maze: time in open arms. C,D. Sucrose preference test: before (C) and following (D) stress procedures. E-H. Nest building assay: nestlet mass before (E) and after stress procedures (F), nest score before (G) and after stress procedures (H). I. Experimental timeline.

